# Per- and Polyfluoroalkyl Substances Exposure in New Jersey Prostate Cancer Survivors: A Pilot Biomonitoring Study

**DOI:** 10.64898/2026.07.08.26357561

**Authors:** Stefanie A. Joseph, Chidinma Opara, Megan R. Shanahan, Julianne Varga, Jennifer Falcon, Uyaiobong Ibanga, Surya Venkatraman, Marley Perlstein, Thomas L. Jang, David Golombos, Saum Ghodoussipour, Tina Fan, Shawn O’Leary, Judith M. Graber, Jaime E. Hart, Emily S. Barrett, Elisa V. Bandera, Hari S. Iyer

**Author notes:** Corresponding Author Hari S. Iyer, ScD MPH Rutgers Cancer Institute, 120 Albany St. Tower 2, Office 8009 New Brunswick, NJ 08901.

## Abstract

**Background:** Men with prostate cancer (PCa) may be especially vulnerable to per- and polyfluoroalkyl substances (PFAS) exposure due to their endocrine-disrupting and cardiometabolic impacts and cardiotoxicity and immune suppression of treatments.

**Objective:** A pilot study was launched to measure serum and tap water PFAS concentrations in PCa survivors.

**Methods:** Men with PCa were recruited from Rutgers Cancer Institute between February 2025 and March 2026, with ongoing enrollment and follow-up. Eligible men were aged ≥40 years on active surveillance or within 3-12 months of initial definitive treatment. Participants provided blood and residential tap water samples, which were analyzed using mass spectrometry (serum) and modified EPA method 537 (water). Geometric means were used to summarize PFAS concentrations by race and assess serum-tap water correlations.

**Results:** Of 235 eligible patients, 124 (60%) enrolled. Median age was 64 years; 63% were non-Hispanic White, 43% had a Gleason score ≤6. Roughly half of participants provided serum and/or tap water samples. In serum, six PFAS analytes had >80% detection; of these analytes, median concentrations ranged from 0.13 ng/mL (IQR: 0.07-0.20) for PFHpS to 2.55 ng/mL (IQR:1.54-3.82) for nPFOS. Among 74 tap water samples, 9 PFAS analytes had >60% detection; of these, median concentrations of PFNA (0.56 ng/L; IQR: 0.33–0.75), PFOA (3.75 ng/L; IQR: 1.21–5.27), and PFOS (2.29 ng/L; IQR: 0.46–2.89), were below New Jersey Maximum Contaminant Levels. Non-White participants had significantly higher levels of multiple PFAS analytes in both serum and tap water. Serum-tap water correlations were modest (r=0.22–0.41).

**Significance:** The pilot study has demonstrated both the feasibility and importance of studying PFAS exposure pathways as well as potential impacts of PFAS exposure in diverse populations.

**Impact Statement:** This study provides some of the first estimates of PFAS exposure among prostate cancer patients in serum and tap water, showing moderate correlations between tap water and serum concentrations of specific PFAS analytes. These findings can support larger studies to identify environmental exposure sources and evaluate the role of PFAS in prostate cancer progression and outcomes.

## Introduction

Per- and polyfluoroalkyl substances (PFAS) are a key class of chemical pollutants of environmental and human health concern due to their ubiquitous presence in water systems and adverse health impacts (1). PFAS are highly persistent in soil, air and water, with those commonly measured in humans having half-lives ranging from 2.7-8.5 years (2). Even at low exposure levels, PFAS can bioaccumulate in human bodies for years. PFAS have also been linked to immune and hormonal disruption, cardiovascular disease (CVD) risk factors primarily dyslipidemia, liver and kidney dysfunction (3,4). Emerging evidence suggests PFAS exposure may increase risks of certain cancers (5). However, there has been limited research on impacts of PFAS and cancer survivors, who may face increased health risks from exposure due to cardiotoxicity of certain treatments (6).

There are approximately 3.5 million men in the United States living with prostate cancer (PCa) accounting for 42% of all male cancer survivors (7,8). Evidence for risks of PCa associated with PFAS is mixed, with some studies finding higher risks in certain subgroups (9), while others find no association (10). Through earlier detection of PCa and advances in treatment, nearly all survivors can expect to live at least 15 years after diagnosis of PCa (8). Because of this, CVD is a major competing risk of death, particularly among men diagnosed with indolent tumors (11). In addition, hormonal androgen deprivation therapy (ADT), a cornerstone of treatment for PCa, may increase CVD risks over long durations (12).

There is growing interest in identifying modifiable factors, including the built and natural environment and pollutants, that may influence long-term CVD and PCa outcomes (13–19). Due to their endocrine-disrupting and cardiometabolic impacts (20), exposure to PFAS may negatively impact treatment and survival outcomes among men with PCa. Men diagnosed with PCa may be particularly susceptible to PFAS exposure through endocrine-disrupting and lipid-related pathways (21,22). PFAS exposure may interfere with anticancer actions of certain drugs used for treatment of castration-resistant PCa, reducing efficacy through pathways involving oxidative stress and protein binding (23). Since PFAS can disrupt androgen receptor signaling, among cancer patients exposure may additionally interfere with the metabolic effects of ADT through synergistic influences on testosterone suppression (22). Therefore, PFAS and ADT may exacerbate cardiometabolic risks in PCa survivors through convergent pathways.

In order to study potential effects of PFAS exposure on PCa progression and cardiometabolic outcomes in survivors, the “Research on the Epidemiology of Prostate Cancer and Environmental Levels” (REPEL) cohort was launched. This pilot study focused on recruiting participants in New Jersey due to the state’s historical challenges with PFAS contamination in drinking water systems (24). Data reported by the Unregulated Contaminant Monitoring Rule 3 from 2013 to 2015 found higher PFAS contamination in New Jersey compared to other states, and higher PFAS levels were correlated with proximity to industrial waste sites (25). Following evidence of state-wide detections of PFAS in water systems and evidence of health risks (26), New Jersey became the first state in the United States to enact Maximum Contaminant Levels (MCLs) for PFAS in drinking water in 2018. Further historical details about PFAS monitoring and regulations in New Jersey are presented in the **Supplementary Methods**. Setting legally-enforced MCLs led to roughly 50% declines in PFOA and PFNA concentrations in New Jersey water systems (27), but New Jersey residents may still experience ongoing PFAS exposure from other routes of exposure or drinking water sources as well as private wells.

Here, we describe the pilot study design and implementation, including recruitment procedures and collection of multimodal data, biospecimens, and environmental samples from participants. We summarize descriptive characteristics and PFAS levels in a sample of individuals with serum and residential tap water analyzed for PFAS. These findings will help to understand PFAS exposure sources and serum levels in a contemporary population of men with PCa.

## Methods

### Study population, eligibility, and recruitment

The REPEL study was conducted at the Rutgers Cancer Institute of New Jersey (CINJ) in New Brunswick. PCa incidence in New Jersey is higher than the national average (142.6 vs. 113.2 per 100,000 men) (28). The New Brunswick CINJ site was chosen because it is centrally located, enabling recruitment of study participants from across New Jersey (**Figure 1**).

**Figure 1.**
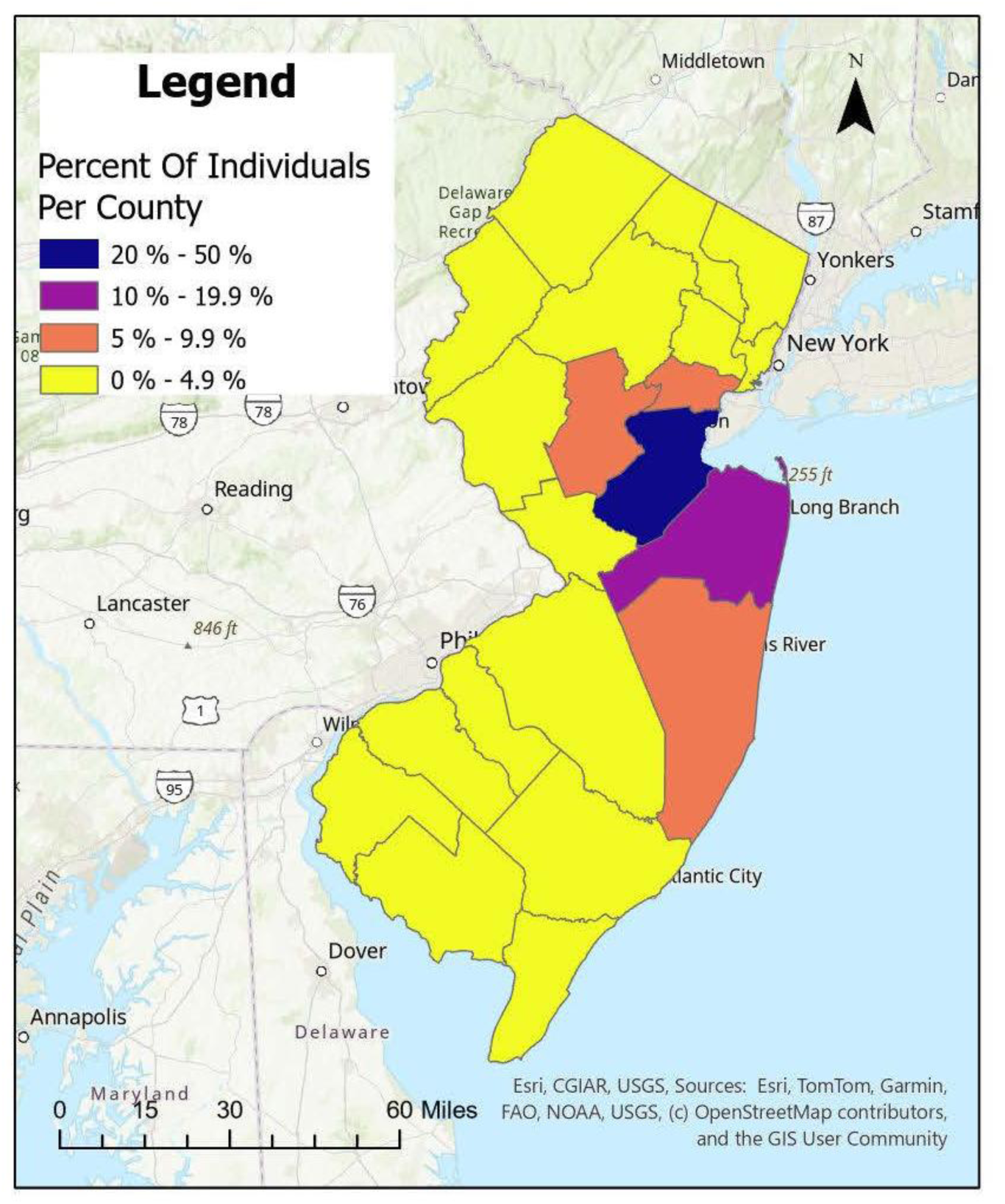
Geographic locations of REPEL study participants with questionnaire data recruited 02/2025 to 03/2026 (N=115)

Eligible men were aged ≥40 years, diagnosed with primary PCa, had a permanent residence in New Jersey, visited CINJ for treatment, and were between 3 and 12 months of initial definitive treatment (radiation, hormone therapy, or surgery) or on active surveillance. This treatment window was selected to limit participant burden during the acute phase of treatment and recovery, and capture men who were most engaged with their care and therefore likely to participate. Men on active surveillance could be enrolled any time after their diagnosis. Patients who experienced cognitive impairment, blindness or deafness or were non-English speakers were excluded from enrollment.

Study staff screened patients for eligibility using the Robert Wood Johnson (RWJ) Barnabas Health EPIC electronic health record (EHR) system. Each week, the REPEL team checked upcoming appointments among patients scheduled to see genitourinary (GU) oncology care providers. For patients who were deemed eligible, REPEL staff notified providers about patients to be approached for study participation. Patients were first introduced to the study by GU oncologists during their visit. If the patient expressed interest, REPEL staff were then invited to meet the patient and administer informed consent.

A waiver of informed consent was obtained to access medical records for screening based on minimal risk to participants and feasibility. The study was approved by the Rutgers University Institutional Review Board. Written informed consent was obtained from participants.

### Data collection

Participants were invited to complete a questionnaire within three weeks of enrollment. The REPEL team coordinated with patients and providers to schedule blood collection at their next clinic visit within 6-12 months of enrollment. For the pilot study, participants were invited to provide tap water samples, and those who completed all study procedures received a $50 gift card as a token of appreciation. The overall study design is presented in **Supplementary Figure 1**. Details about stakeholder engagement and the Community Advisory Board (CAB) are described in **Supplementary Figure 2** and **Supplementary Methods**.

### Questionnaires

The REPEL questionnaire was developed using prior instruments for PFAS exposure assessment (29) and captured information on demographics, education and income, and smoking status. Residential history was collected to inform future modeling of PFAS exposure across the life course and support subsequent geospatial analyses and linkages (30). Questions on PFAS exposure assessment were adapted from established PFAS exposure assessment tools and validated dietary instruments used in prior studies (29,31–35), and captured information on home water filters and drinking water sources, and dietary habits over the past 12 months were also captured to allow for investigation of major sources of PFAS exposure in PCa survivors. Since water and diet are major sources of PFAS exposure, participants were asked detailed questions about use and intake to assess consumption and allow for adjustment in statistical models.

The survey also captured information about any PCa treatment and diagnostic work up received outside of the RWJ Barnabas Healthcare system, or care that occurred prior to use of EPIC at Rutgers Cancer Institute in 2022. Additionally, information about PCa care and management were collected, as well as risk factors including family history of cancer, PCa testing and screening, and PCa recurrence. In order to enable further study on other important health conditions in prostate cancer survivors, we also asked participants about quality of life such as frequency of urinary symptoms within the last four weeks (36), and any treatments, medications, or procedures related to PCa, CVD, kidney, and liver related conditions (37). PCa–related questions and additional health conditions, treatment, and medication items were based on previously developed instruments (38,39). In order to facilitate research on long-term cardiovascular and cardiometabolic diseases, we added questions about these conditions (e.g. diabetes, congestive heart failure) and treatment (39).

The questionnaire was administered virtually via Rutgers secured Zoom platform or by telephone. Survey responses were recorded in a REDCap database. Questionnaires took an average of 40 (SD: 18.6) minutes to complete.

### Medical records extraction

Clinical and treatment information was abstracted from GU oncologists’ progress notes, previously logged encounters, pathology, imaging, laboratory tests, and current medication lists in the EHR. These sources were used to gather information on PCa tumor staging, prostate-specific antigen (PSA) levels over time, and grading. Dates of diagnosis and recurrence, number of cancer tumors, clinical and pathological grading and TNM staging and diagnosis were also recorded. This instrument was adapted from the Surveillance, Epidemiology, and End Results (SEER) Program Coding and Staging Manual (40). Additionally, the start date and end date of any type of PCa treatment (e.g. surgery, chemotherapy, radiation therapy, etc.), treatment status (e.g. treatment given, active surveillance, etc.), SEER surgery code, radiation treatment modality phase, oral medications, and hormone injections, implants, or infusions were captured. To confirm missing or incomplete clinical data, GU oncology care providers reviewed records with study staff (coauthors: TLJ, DG, SG).

### Serum collection and laboratory assays

Serum specimens were collected once from patients at their follow-up appointments, which occurred between 14-15 weeks following their enrollment in the study. Blood orders were placed by each patient’s GU oncologist, and non-fasting samples were drawn on-site at the Rutgers Cancer Institute laboratory by phlebotomists either before or after their clinic visit. Serum was collected in 10 mL red tube tops, delivered by the Rutgers Cancer Institute Biospecimen Repository and Histopathology Service Shared Resource. Up to four vials of blood were obtained per patient. Following collection and post clotting period, blood samples were centrifuged at 1,000g for 15 minutes. Some samples required two to three centrifugation cycles to achieve complete separation. The serum was then aliquoted and stored in polypropylene or polyethylene containers at −70 ± 3 °C. Batch shipments were then transported in 1 mL cryovials in an insulated shipper box with dry ice by courier to the New Jersey Department of Health-Public Health and Environmental Laboratories (NJDOH-PHEL) for PFAS analysis. Then, NJDOH-PHEL’s Environmental Chemical Laboratory Services (ECLS) Chemical Terrorism (CT) laboratory assayed PFAS levels using NJDOH’s PFAS testing method #6304.04, informed by CDC QA/QC procedures and used for biomonitoring population-based studies and occupational studies in New Jersey (41,42). The method utilizes a high-throughput online solid phase extraction system and tandem mass spectrometer to detect 19 target PFAS analytes as low as <0.1 ng/mL, including PFOA, PFOS, and PFNA. Further details about the method have been described previously (42).

### Tap water collection and laboratory assays

Study participants received a tap water collection kit in-person at their scheduled appointment at CINJ. Tap water kits were prepared at CINJ by study staff, with field collection bottles containing Trizma as a preservation reagent for buffering and to remove chlorine. Participants were instructed to fill each of the three sample bottles to the shoulder. Using participant-led collection reduced time and costs associated with professional or volunteer collectors and decreased participant burden by allowing samples to be collected at their convenience. Written instructions, a tap water collection form, and a QR code to a tutorial video were included. When collecting water, participants flushed tap water for 3-5 minutes to achieve a stable stream and temperature. Using gloves to reduce contamination, they collected water in sample and control bottles, agitated them by hand, and kept chilled before delivering them in the kit to UPS for shipping. A prepaid shipping label addressed to NJDOH-PHEL was provided so participants could ship the samples directly to the lab after collection. Kits were distributed between 0.3-10 months from enrollment.

Sample analyses were conducted following protocols outlined in EPA Method 537 (43) to detect 25 PFAS analytes, modified to allow participant self-collection. The method uses solid phase extraction to reconstitute PFAS in samples and liquid chromatography-tandem mass spectrometry (LC-MS/MS) for analysis, with further details available elsewhere (44). We note that because the study used volunteer collection, not trained staff, the method is not a certifiable EPA Method 537. Samples were received by NJDOH-PHEL on average 9 days following kit delivery to participants. Materials included in the kits are presented in **Supplementary Figure 3**. Further details about the tap water collection and other study procedures are available in the **Supplementary Methods**.

### Statistical analysis

Concentrations of PFAS in serum and water were examined using median and IQR to assess measure of central tendency and spread, and arithmetic means (AM), geometric means (GM), and 95% CI to compare to other studies and for comparisons across population subgroups. The frequency of detected PFAS was calculated for individuals with both available serum and tap water PFAS concentrations.

Correlations between tap water and serum PFAS levels were estimate using Spearman correlations. Due to limited sample sizes, racial categories were analyzed without further stratification by ethnicity; the full race and ethnicity distribution is presented in **Table 1**. Prostate cancer risk groups were defined using the National Comprehensive Cancer Network (NCCN) guidelines (45).

**Table 1:**
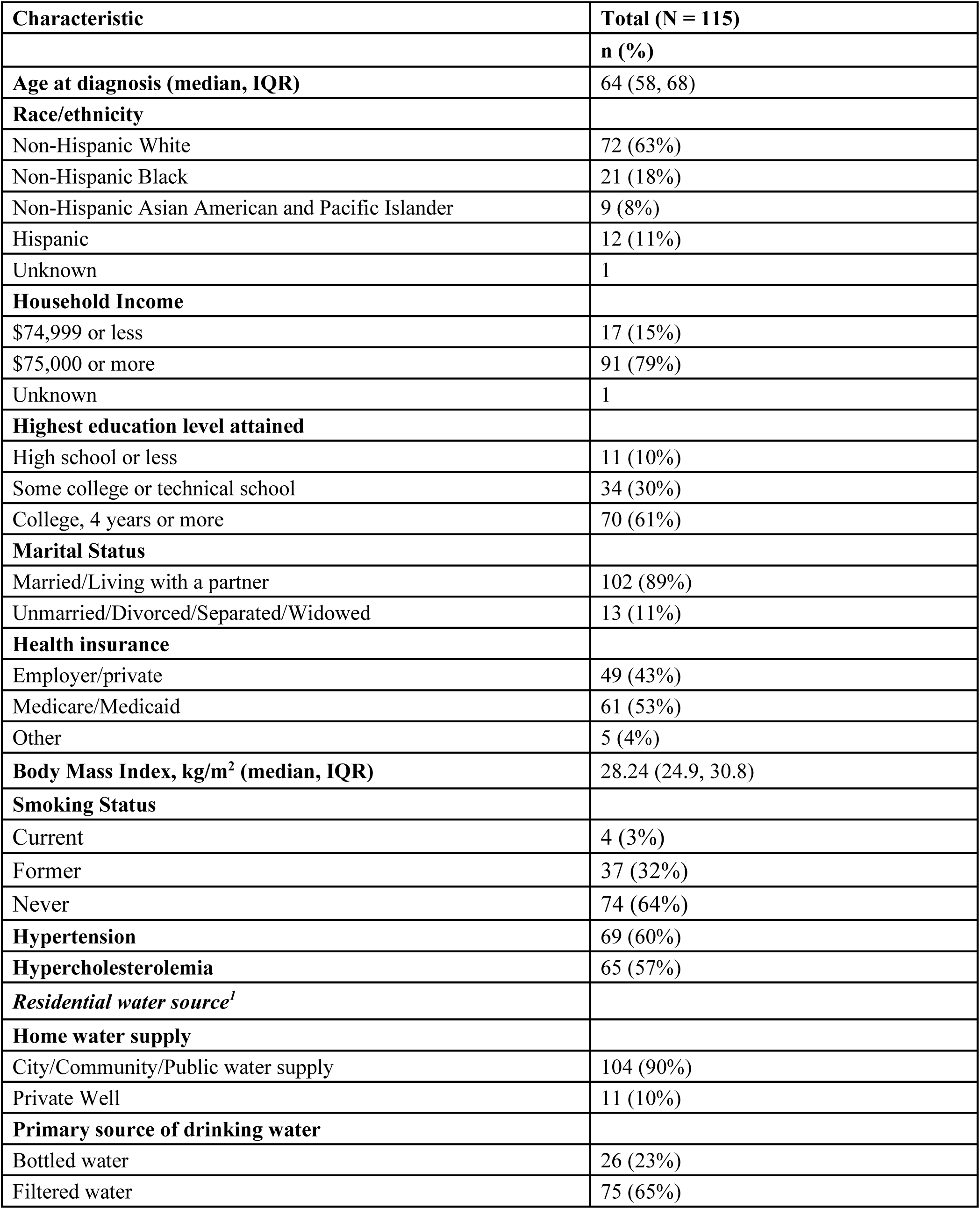
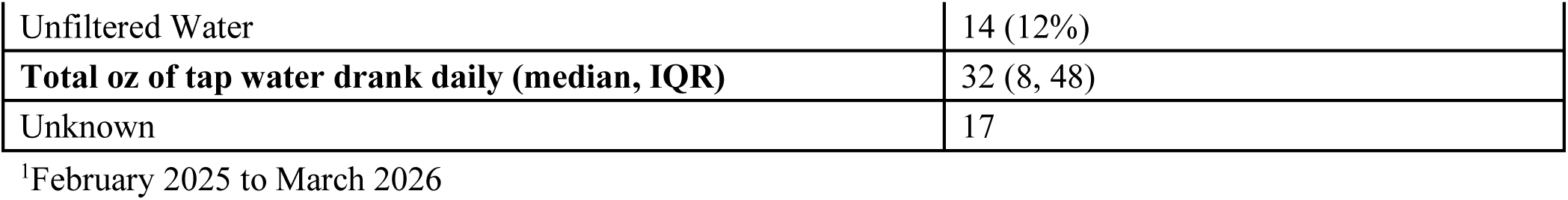
Sociodemographic characteristics of REPEL study participants with questionnaire data, 02/2025 to 03/2026.

| **Characteristic** | **Total (N = 115)** |
| --- | --- |
|  | **n (%)** |
| **Age at diagnosis (median, IQR)** | 64 (58, 68) |
| **Race/ethnicity** |  |
| Non-Hispanic White | 72 (63%) |
| Non-Hispanic Black | 21 (18%) |
| Non-Hispanic Asian American and Pacific Islander | 9 (8%) |
| Hispanic | 12 (11%) |
| Unknown | 1 |
| **Household Income** |  |
| \$74,999 or less | 17 (15%) |
| \$75,000 or more | 91 (79%) |
| Unknown | 1 |
| **Highest education level attained** |  |
| High school or less | 11 (10%) |
| Some college or technical school | 34 (30%) |
| College, 4 years or more | 70 (61%) |
| **Marital Status** |  |
| Married/Living with a partner | 102 (89%) |
| Unmarried/Divorced/Separated/Widowed | 13 (11%) |
| **Health insurance** |  |
| Employer/private | 49 (43%) |
| Medicare/Medicaid | 61 (53%) |
| Other | 5 (4%) |
| **Body Mass Index, kg/m<sup>2</sup> (median, IQR)** | 28.24 (24.9, 30.8) |
| **Smoking Status** |  |
| Current | 4 (3%) |
| Former | 37 (32%) |
| Never | 74 (64%) |
| **Hypertension** | 69 (60%) |
| **Hypercholesterolemia** | 65 (57%) |
| ***Residential water source<sup>1</sup>*** |  |
| **Home water supply** |  |
| City/Community/Public water supply | 104 (90%) |
| Private Well | 11 (10%) |
| **Primary source of drinking water** |  |
| Bottled water | 26 (23%) |
| Filtered water | 75 (65%) |
| Unfiltered Water | 14 (12%) |
| **Total oz of tap water drank daily (median, IQR)** | 32 (8, 48) |
| Unknown | 17 |

<sup>1</sup>February 2025 to March 2026

## Results

### REPEL study recruitment

Over the study period, 235 patients were determined to be eligible after EHR screening. REPEL staff notified providers to approach these patients for potential study enrollment, of which 207 were seen. Overall, 124 (60%) patients who were approached by the REPEL staff agreed to participate **(Supplementary Figure 4)**. Complete questionnaire data was available for 115 participants. At the time of analysis, 74 tap water samples and 63 serum samples were collected. **Supplementary Table 1** provides descriptive characteristics of participants stratified by enrollment status, demonstrating that characteristics are similar between the two groups.

### Cohort sociodemographic and prostate cancer characteristics

**Table 1** presents the characteristics of those who completed the questionnaire. The study recruited men with a median [IQR] age of 64 [58-68 years], who were mostly non-Hispanic White (63%) and married/living with a partner (89%). Over half reported being diagnosed with hypertension (60%) and hypercholesterolemia (57%). PCa tumor characteristics and risk factors are presented in **Table 2**. Family history of any cancer was reported in 25% of participants. Most participants were classified as NCCN low risk (34%) or intermediate risk (41%). There were 36 (31%) men who underwent a radical prostatectomy, 24 (21%) who received ADT, and 12 (10%) men received radiation therapy. Most men reported no or few instances of PCa symptoms in the past four weeks.

**Table 2:**
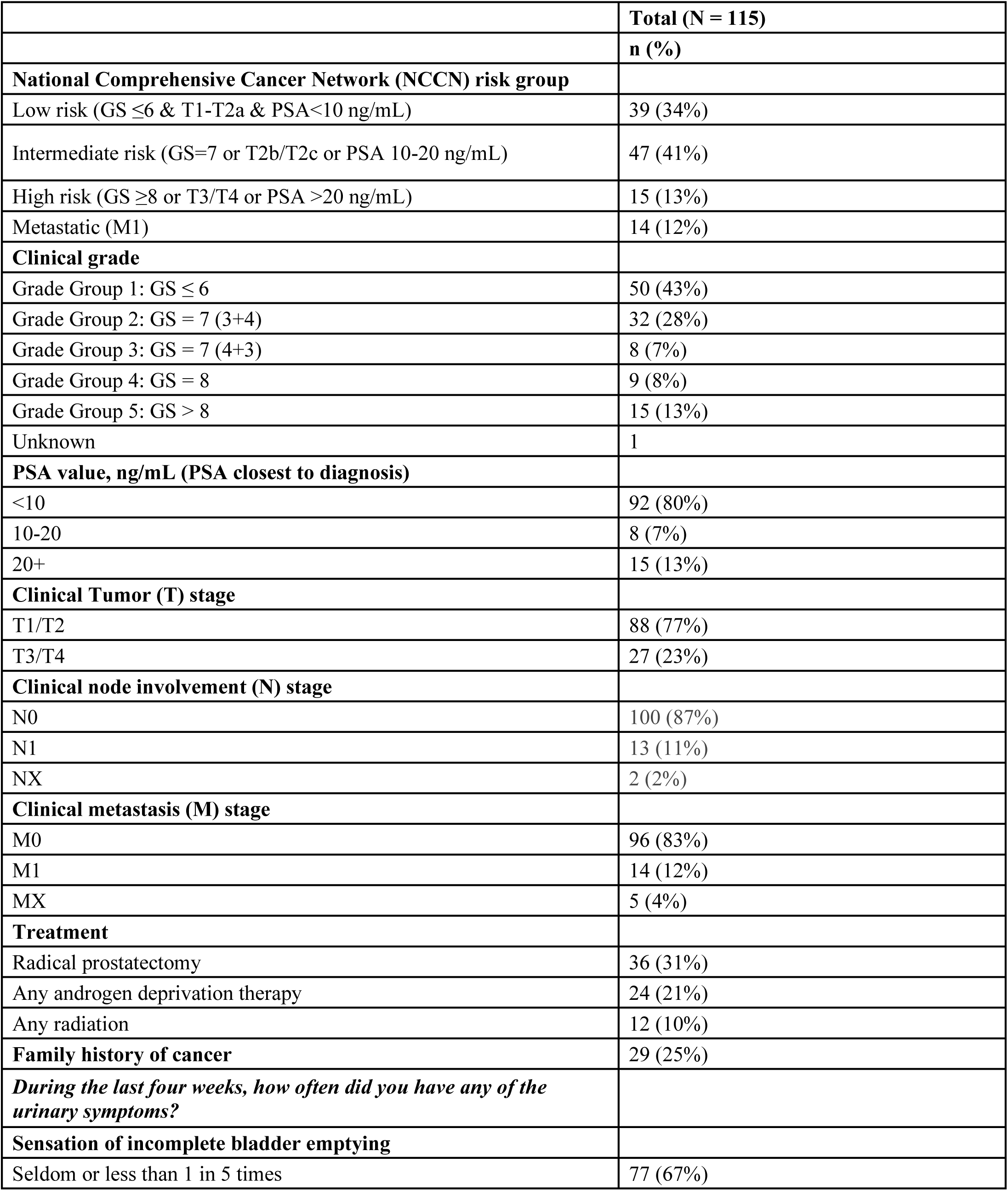
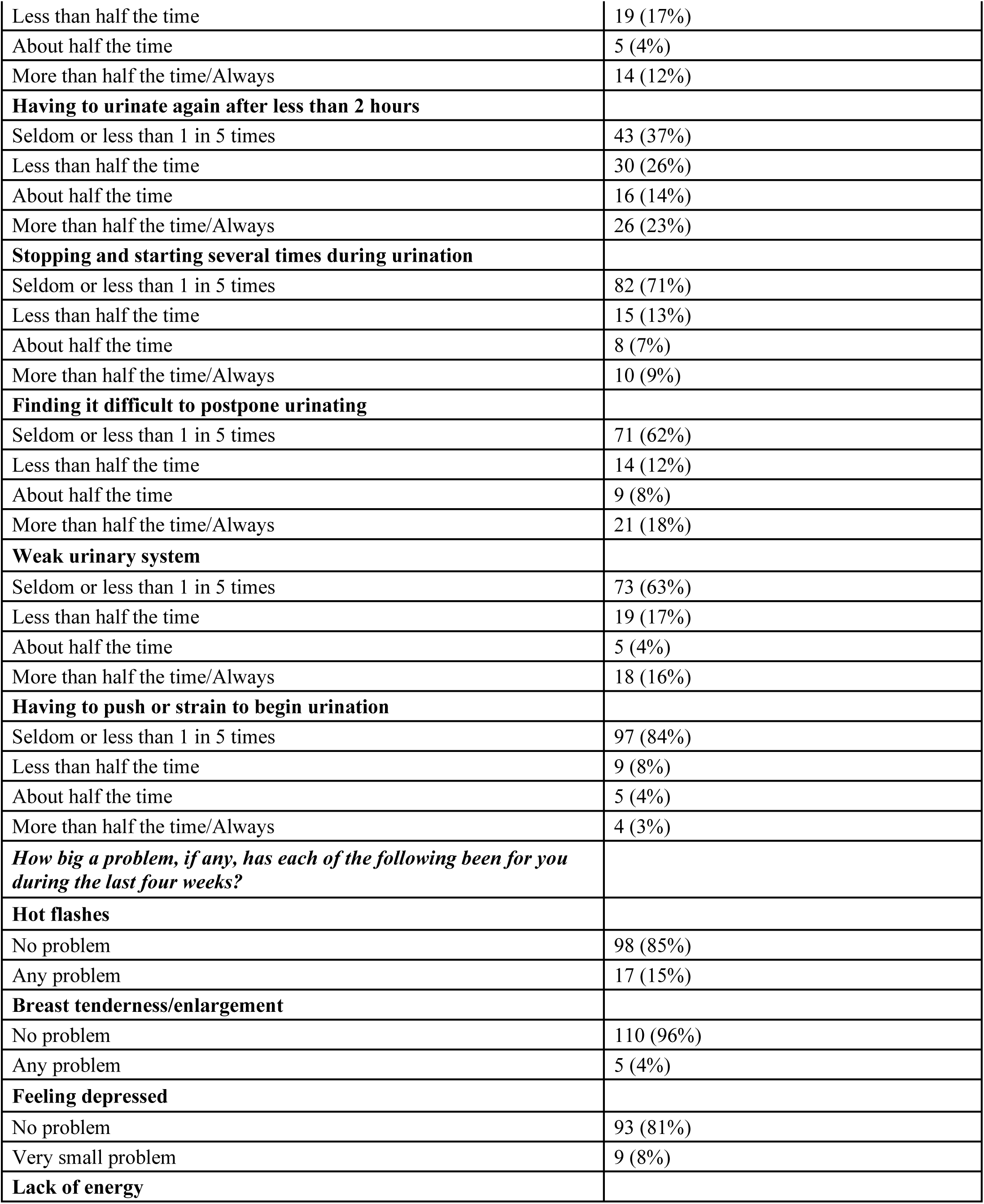
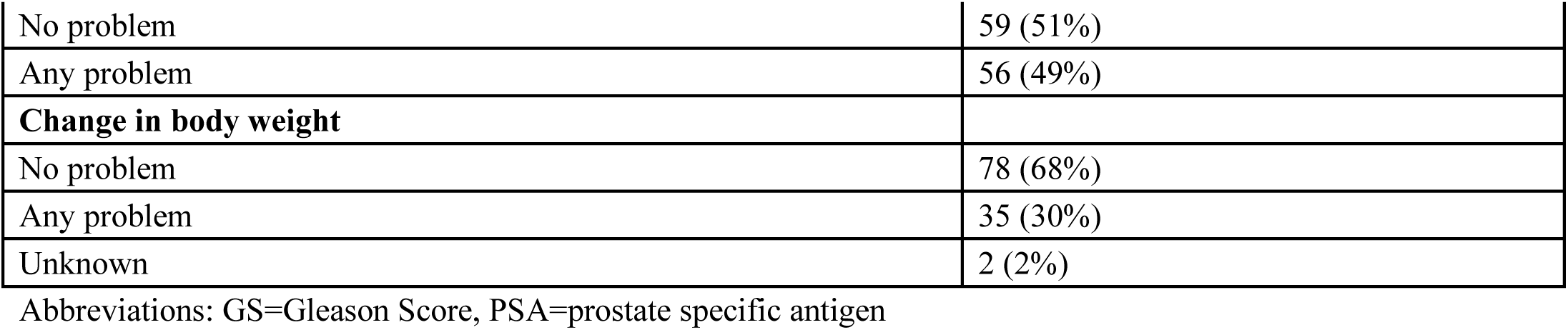
Prostate cancer tumor, treatment and quality of life among REPEL study participants with questionnaire data, 02/2025 to 03/2026.

|  | **Total (N = 115)** |
| --- | --- |
|  | **n (%)** |
| **National Comprehensive Cancer Network (NCCN) risk group** |  |
| Low risk (GS ≤6 & T1-T2a & PSA <10 ng/mL) | 39 (34%) |
| Intermediate risk (GS=7 or T2b/T2c or PSA 10-20 ng/mL) | 47 (41%) |
| High risk (GS ≥8 or T3/T4 or PSA >20 ng/mL) | 15 (13%) |
| Metastatic (M1) | 14 (12%) |
| **Clinical grade** |  |
| Grade Group 1: GS ≤ 6 | 50 (43%) |
| Grade Group 2: GS = 7 (3+4) | 32 (28%) |
| Grade Group 3: GS = 7 (4+3) | 8 (7%) |
| Grade Group 4: GS = 8 | 9 (8%) |
| Grade Group 5: GS > 8 | 15 (13%) |
| Unknown | 1 |
| **PSA value, ng/mL (PSA closest to diagnosis)** |  |
| <10 | 92 (80%) |
| 10-20 | 8 (7%) |
| 20+ | 15 (13%) |
| **Clinical Tumor (T) stage** |  |
| T1/T2 | 88 (77%) |
| T3/T4 | 27 (23%) |
| **Clinical node involvement (N) stage** |  |
| N0 | 100 (87%) |
| N1 | 13 (11%) |
| NX | 2 (2%) |
| **Clinical metastasis (M) stage** |  |
| M0 | 96 (83%) |
| M1 | 14 (12%) |
| MX | 5 (4%) |
| **Treatment** |  |
| Radical prostatectomy | 36 (31%) |
| Any androgen deprivation therapy | 24 (21%) |
| Any radiation | 12 (10%) |
| **Family history of cancer** | 29 (25%) |
| *During the last four weeks, how often did you have any of the urinary symptoms?* |  |
| **Sensation of incomplete bladder emptying** |  |
| Seldom or less than 1 in 5 times | 77 (67%) |
| Less than half the time | 19 (17%) |
| About half the time | 5 (4%) |
| More than half the time/Always | 14 (12%) |
| **Having to urinate again after less than 2 hours** |  |
| Seldom or less than 1 in 5 times | 43 (37%) |
| Less than half the time | 30 (26%) |
| About half the time | 16 (14%) |
| More than half the time/Always | 26 (23%) |
| **Stopping and starting several times during urination** |  |
| Seldom or less than 1 in 5 times | 82 (71%) |
| Less than half the time | 15 (13%) |
| About half the time | 8 (7%) |
| More than half the time/Always | 10 (9%) |
| **Finding it difficult to postpone urinating** |  |
| Seldom or less than 1 in 5 times | 71 (62%) |
| Less than half the time | 14 (12%) |
| About half the time | 9 (8%) |
| More than half the time/Always | 21 (18%) |
| **Weak urinary system** |  |
| Seldom or less than 1 in 5 times | 73 (63%) |
| Less than half the time | 19 (17%) |
| About half the time | 5 (4%) |
| More than half the time/Always | 18 (16%) |
| **Having to push or strain to begin urination** |  |
| Seldom or less than 1 in 5 times | 97 (84%) |
| Less than half the time | 9 (8%) |
| About half the time | 5 (4%) |
| More than half the time/Always | 4 (3%) |
| ***How big a problem, if any, has each of the following been for you during the last four weeks?*** |  |
| **Hot flashes** |  |
| No problem | 98 (85%) |
| Any problem | 17 (15%) |
| **Breast tenderness/enlargement** |  |
| No problem | 110 (96%) |
| Any problem | 5 (4%) |
| **Feeling depressed** |  |
| No problem | 93 (81%) |
| Very small problem | 9 (8%) |
| **Lack of energy** |  |
| No problem | 59 (51%) |
| Any problem | 56 (49%) |
| **Change in body weight** |  |
| No problem | 78 (68%) |
| Any problem | 35 (30%) |
| Unknown | 2 (2%) |

Abbreviations: GS=Gleason Score, PSA=prostate specific antigen

### Dietary exposures and water consumption

Food and drink sources were documented during the dietary habit section of the questionnaire to assess possible sources of PFAS exposure. Of the specific food items queried with links to PFAS, the most commonly consumed were meat, poultry, fish, or cold cuts wrapped in plastic, followed by water in plastic containers. Most men reported filtered water (65%) as their primary source (**Table 1**). Further details are provided in **Supplementary Results**, **Supplementary Table 2, and Supplementary Figure 5**.

### Tap water PFAS concentrations

Twenty-five PFAS analytes were measured in tap water samples collected by participants from their homes. **Supplementary Table 3** lists summaries of analytes evaluated in water collected by 74 participants. **Figure 2** shows analytes present in tap water with a regulated New Jersey MCL and U.S. Environmental Protection Agency (EPA) guideline. Of these, 9 had detectable levels in >60% of individuals, with median [IQR] ranging from 0.56 [0.33-0.75] ng/L in analyte PFNA to 3.75 [1.21-5.27] ng/L in analyte PFOA. Overall, levels were lower than New Jersey PFAS MCL regulations set for PFOA, PFOS and PFNA (13-14 ng/L), but the 90^th^ percentile for PFOS (4.52 ng/L) and PFOA (8.04 ng/L) exceeded EPA’s PFAS MCLs (4 ng/L) proposed in 2024 (46).

**Figure 2:**
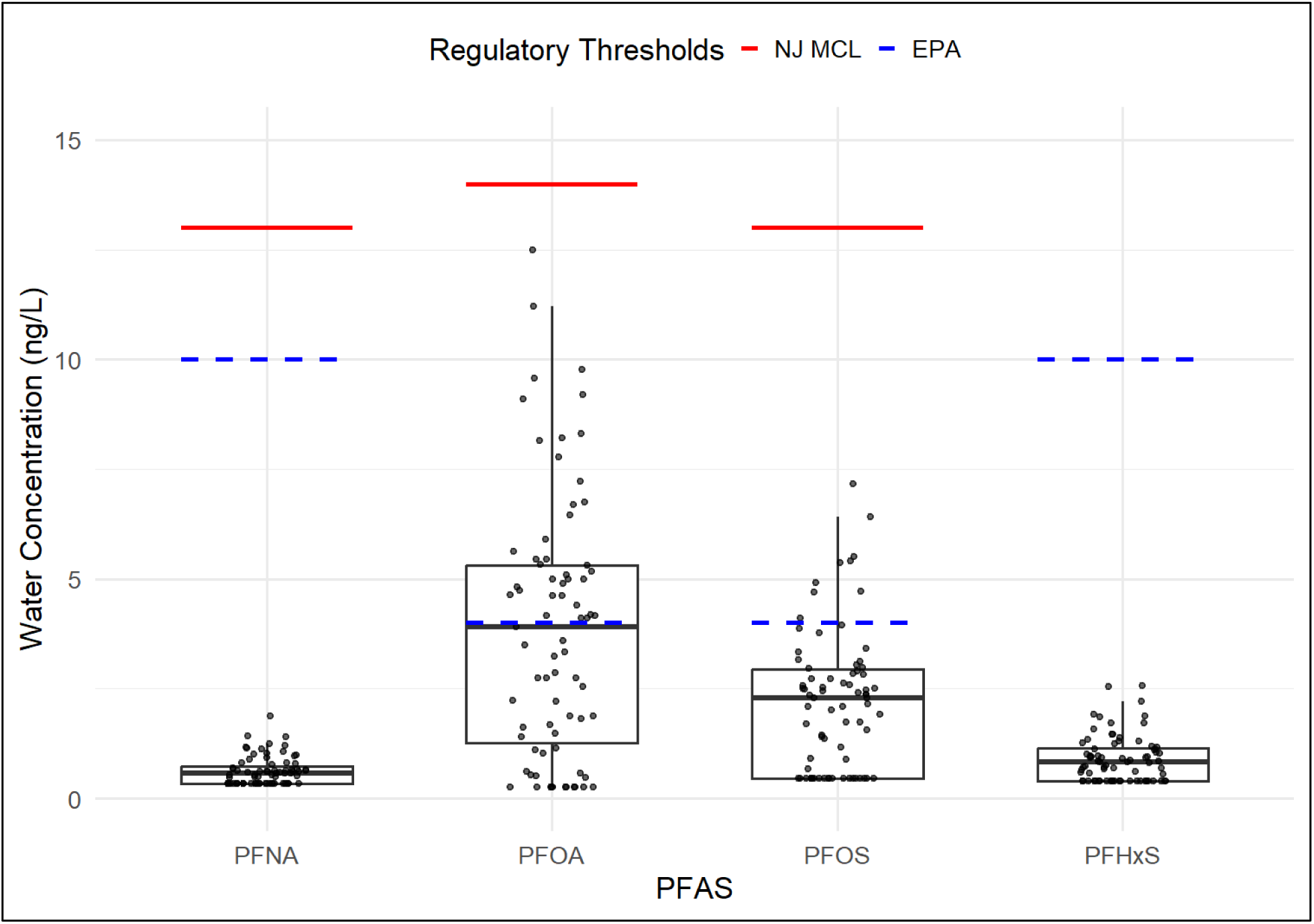
PFAS levels in residential tap water from REPEL participants (N=74) for analytes with New Jersey and EPA regulatory limits in drinking water

We compared tap water PFAS levels by age, race, and NCCN risk group (**Figure 3, Supplementary Table 4**). Compared to White participants, non-White participants had higher tap water concentrations of PFHxS.

**Figure 3:**
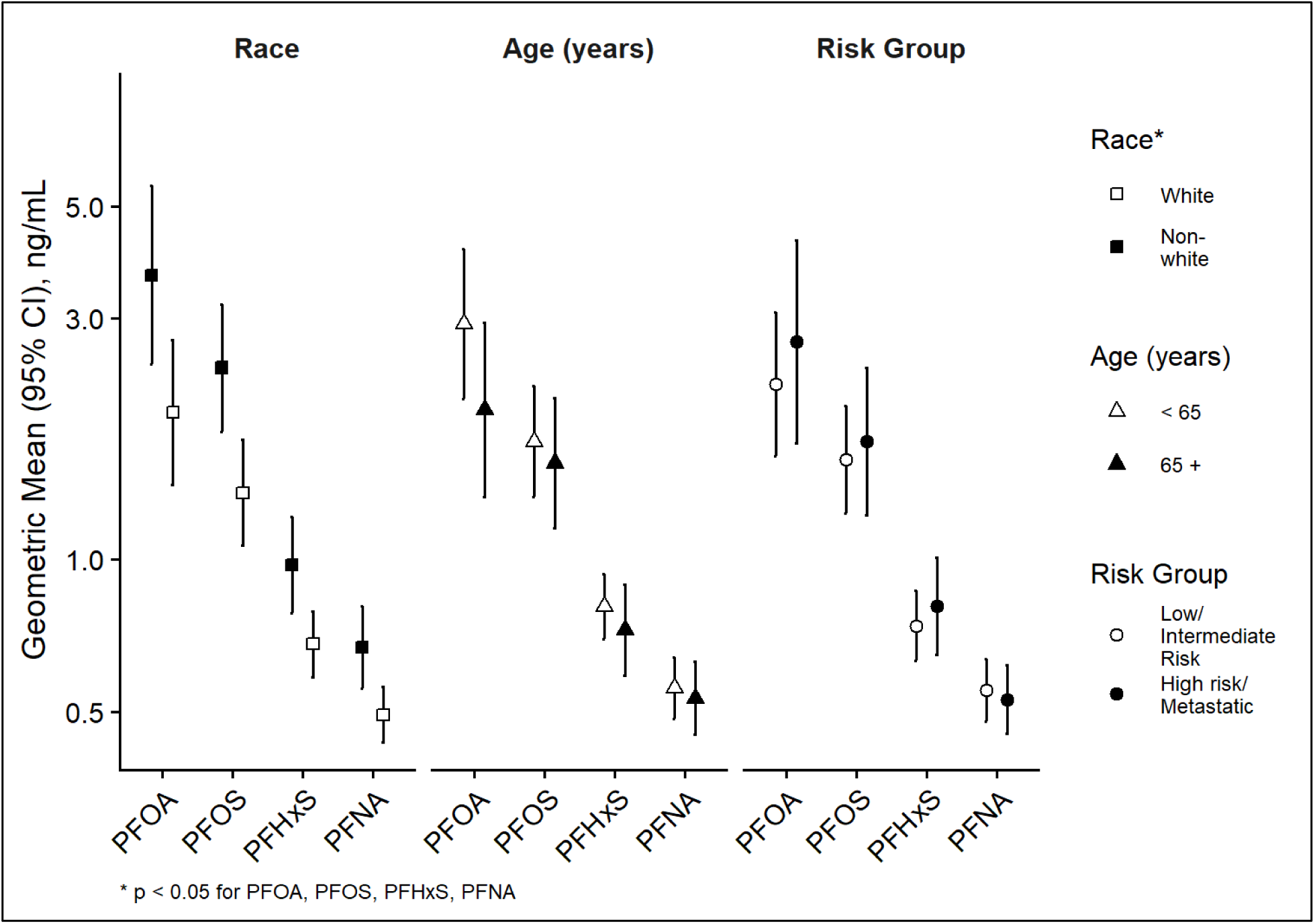
Concentration of PFAS in residential tap samples of participants (N=74) by age, race, and risk group among four analytes with NJ MCL and EPA regulations. Note: Due to limited sample sizes, racial categories were analyzed without further stratification by ethnicity.

### Serum PFAS concentrations

Nineteen analytes were assessed in serum for 63 participants with available data **(Supplementary Table 5)**; Of these, nine PFAS were detected in >50% of participants, with median concentrations ranging from non-detect to 2.55 ng/mL. Maximum concentrations exceeded 10 ng/mL for smPFOS (12.30 ng/mL) and nPFOS (30.70 ng/mL).

PFAS with ≥80% detection are shown in **Figure 4** and **Supplementary Table 4** by age, race, and NCCN risk group. As with tap water, significant differences were observed by race for PFHpS. Compared to NCCN low/intermediate risk participants, those with high risk/metastatic disease had higher serum levels for all six PFAS analytes, though these differences were not statistically significant.

**Figure 4:**
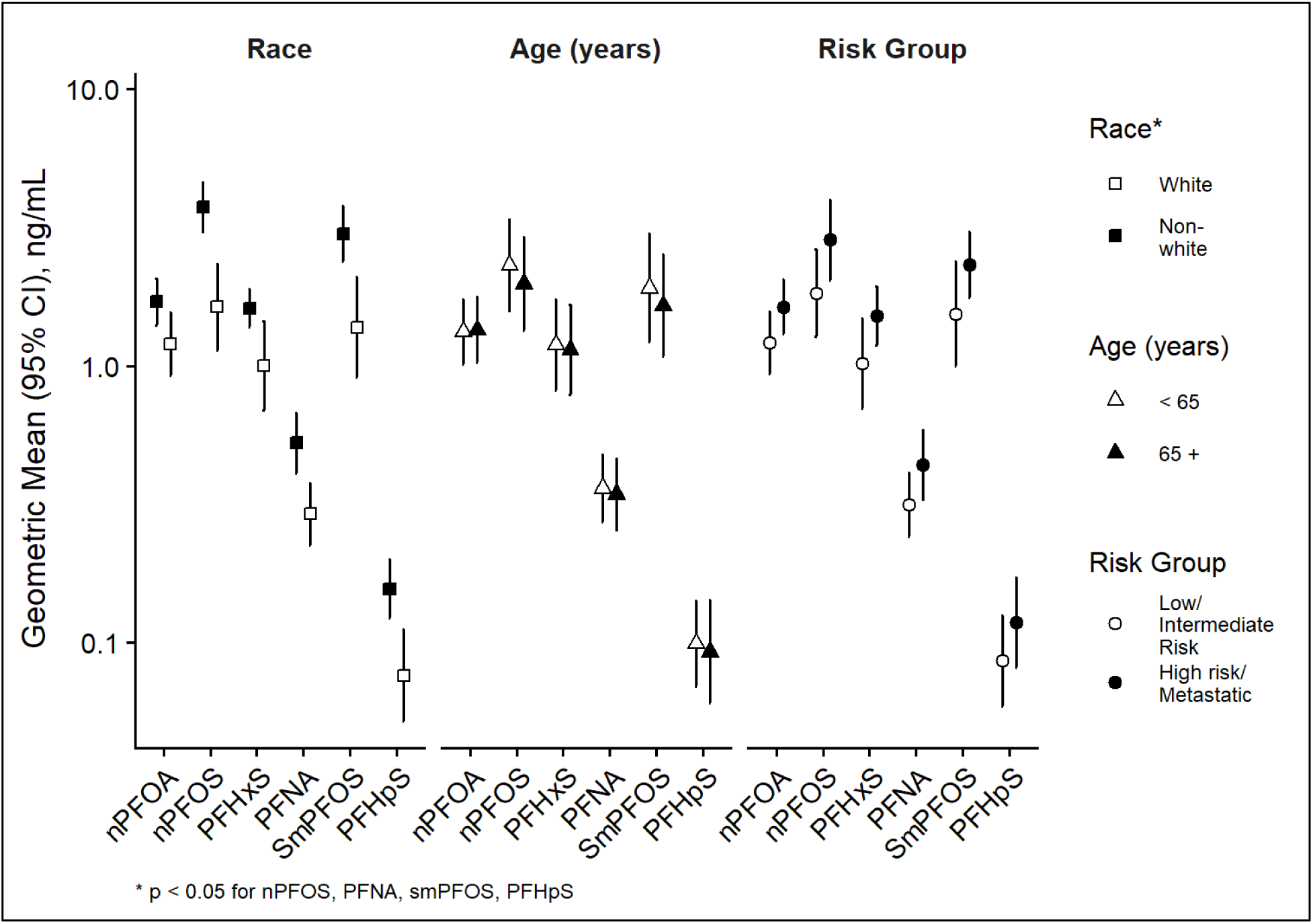
Concentration of PFAS in serum samples of participants (N=63) by age, race, and risk group among six analytes with 80% detection in serum. Note: Due to limited sample sizes, racial categories were analyzed without further stratification by ethnicity.

### Correlation between PFAS in tap water and PFAS in serum

Fifty-four participants provided both tap water and serum samples. Correlations between the four PFAS with state and federal regulatory limits in tap water and serum were examined (**Figure 5**). There was low to moderate correlation between tap water and serum levels within each analyte, ranging from 0.22 and 0.41. Strong correlations (r=0.60-0.80; *P* <0.001) were observed among PFAS analytes within serum samples. Correlations between PFAS analytes appeared higher (r ≥ 0.80; *P* <0.001) within tap water samples.

**Figure 5:**
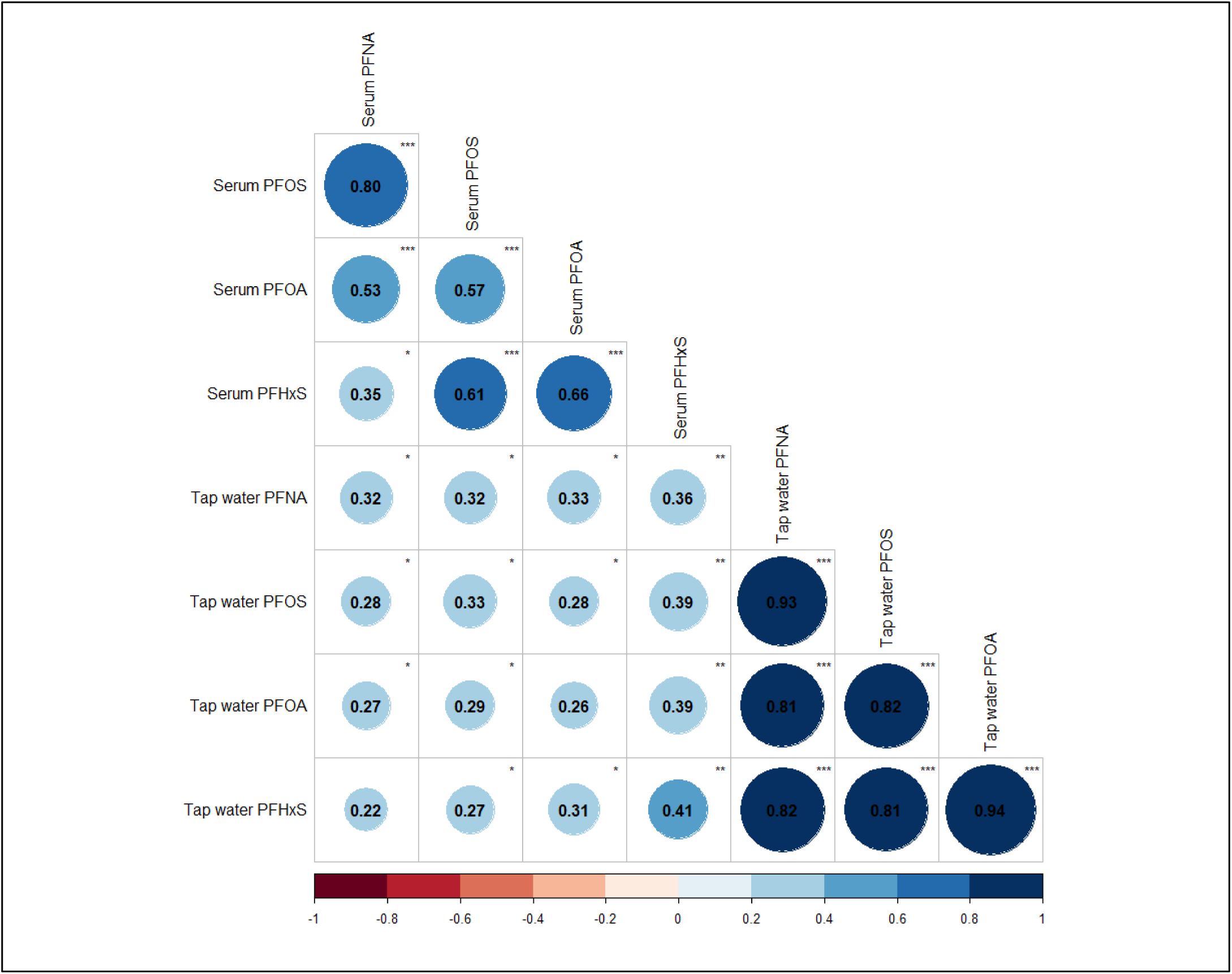
Correlation matrixes between PFAS in residential tap water and PFAS in serum (N=54) Legend: *p<0.05, **p<0.01, ***p<0.001

## Discussion

The REPEL pilot study is the first to examine PFAS levels among men with PCa in New Jersey. Consistent with other studies serum concentrations of PFOA and PFOS were higher compared to the other PFAS analytes, perhaps reflecting their long use for manufacturing in the U.S. (47,48). Correlations between PFAS analytes were stronger within the same medium than across water and serum samples. The modest correlations between water and serum PFAS could be due to temporal misalignment, with water reflecting current PFAS exposure levels, while serum reflecting chronic and longer-term burden of PFAS (49), or that other exposures are driving serum levels. We observed exposure-related racial disparities, with non-White individuals having higher average tap water and serum concentrations of PFAS than White individuals in the study.

In REPEL, drinking water emerged as potentially important source of PFAS exposure in this population of New Jersey PCa survivors. A high reported intake of bottled water among participants was found and the detection of PFAS in tap water samples, although concentrations were below New Jersey regulatory limits. Our study provides a contemporary assessment of tap water concentrations in a New Jersey-based sample of elderly men, and showed detections of PFHxA, PFOA, and PFBA in over four out of five samples. Concentrations of tap water PFAS analytes were higher in REPEL than in a study conducted in Indiana households in 2020, where no samples exceeded EPA’s MCLs of 4.0 ng/L for PFOA or PFOS (50). A nation-wide assessment of tap water samples collected from residence, businesses, and drinking water treatment plants from 2016 to 2021 found higher median concentrations of PFOS (4.2-4.3 ng/L), but lower median concentrations of PFOA (2.6-2.8 ng/L) than in REPEL (51). The authors noted that in addition to PFOA, PFBS and PFHxS were the most commonly detected PFAS across laboratories, and that EPA MCLs for PFOA and PFOS were exceeded in 6.7% and 4.2% of samples. Detection frequencies were higher in REPEL, possibly due to higher levels of historic contamination in New Jersey.

Our study also provides examinations of correlations between individual tap water and serum concentrations of PFAS in a more recent sample (2025-2026) than earlier studies. Individual-level correlations of tap water and serum PFAS were modest and could either represent a lower contribution of drinking water to serum levels, or a mismatch of contemporary water sampling vs chronic exposure captured in serum. Nonetheless our study is consistent with others that have shown correlations of drinking water and serum PFAS measures. A recent study conducted in southern Michigan evaluated PFAS in both tap water and blood serum among residents who consumed tap water and lived near a paper mill landfill between 2005 and 2018 (52). Samples collected between 2020 and 2021 showed residents with municipal wells had higher levels of PFOA, PFOS, and PFHXs in their unfiltered tap water. These analytes were also the most abundant in serum samples. Participants with higher PFAS concentrations in their tap water had serum concentrations that were approximately 15-fold higher for PFOA, 5-fold higher for PFOS, and 2-fold higher for PFHxS compared with participants whose tap water contained lower PFAS levels. In Gustavus, Alaska, researchers analyzed 25 residential drinking water samples from private wells located near an airport and fire-training sites, along with 40 blood samples from local residents. PFOS and PFHxS were approximately 70% of the PFAS detected in both serum and water samples. The study identified a positive correlation between PFAS levels in drinking water and serum concentrations, suggesting that drinking water exposure is a major contributor to PFAS bioaccumulation in humans (53). A study of women in the California Teachers Study found 38% higher median serum levels of PFOA and 29% higher median serum levels of PFOS among those residing in areas with detectable levels of these analytes in water systems serving their area using geospatially-derived estimates of PFAS (54). The California Regional Exposure study found roughly 30% higher serum levels of PFOA and PFOS among those residing in areas where drinking water systems had detectable levels of these analytes (55). A study in North Carolina found no association between serum and drinking water and suggested that a temporal mismatch between concentrations in water vs serum could explain this finding (56).

To contextualize PFAS serum concentrations in the REPEL cohort, we compared our findings with those reported in U.S. population-based studies, highly exposed cohorts, and prostate cancer survivor cohorts with PFAS-contaminated drinking water, identified from previous regulatory reports **(Supplementary Figure 6)** (57). REPEL levels were similar to those reported in the New Jersey Health and Nutrition Examination Survey (NJHANES) 2016-2018 (42) and NHANES 2017-2020 (49) for PFOS, PFOA, PFHxS. However, data from NHANES 2005-2014 (58) in working U.S. men aged 20 to 65+ years old showed higher levels than more recent U.S. population based cohorts, potentially reflecting secular declines in PFAS exposure over time. In contrast, serum PFAS concentrations collected in 1993 and 2001 in the Prostate, Lung, Colon, and Ovary Prospective Study (PLCO) cohort (10), cases of 750 men with aggressive PCa were higher than those observed in the other population-based cohorts, except for PFNA. Men without PCa in the PLCO controls group (10) had similar PFAS concentrations to cases. In highly exposed populations, GM PFOS and PFOA serum concentrations generally exceeded 10 ng/mL, with the exception of the levels reported among men and women in Portsmouth, New Hampshire (2019-2021) (59) and New Jersey adults (2019-2023) (60). Mean PFOS levels above 30 ng/mL were observed in men and women in East Metro, Minnesota (2008-2009) (61) and Decatur, Alabama (2010) (62), and in PFOA levels among 60-69 year old men in Hoosick Falls, NY (2016) (63) and men ≥60 years old in the C8 Health Project study (2005-2006) (64).

Racial disparities in PFAS tap water and serum exposure observed in the REPEL study are supported by some but not all prior studies. Although earlier studies have revealed that racial disparities in PCa are driven in part by neighborhood built and social environments (19), there has been more limited research on environmental pollutants as potential mechanisms through which neighborhood social deprivation leads to PCa disparities (16,65). Studies have shown public water systems in the United States serving Hispanic/Latin and non-Hispanic Black residents have higher levels of PFAS contamination in their drinking water across the United States and in New Jersey (26,66,67). Researchers in California have also found higher levels of pesticide application and PFAS near drinking water wells close to communities of racial and ethnic minority groups (67). Other findings have shown higher serum PFAS concentrations in White individuals compared with other racial groups (68–70). Consistent with these findings, an analyses of NHANES serum samples collected between 2011-2018 reported higher concentrations of PFOA in non-Hispanic Whites than Mexican Americans (71). The same study also found that non-Hispanic Whites had higher levels of PFHxS in their serum compared to other racial and ethnic groups (71). Recent studies of PFAS concentrations across racial groups point to specific consumer product use and dietary patterns, including certain dental, hair, and fast food products that may explain different patterns of disparities in different samples (68). Larger studies are needed to confirm this finding.

The REPEL study focuses on PCa survivorship, allowing for longitudinal assessment of key biological pathways of PCa progression and cardiometabolic dysregulation that may impact survivorship outcomes. PFAS may affect PCa outcomes and progression through sex hormone-dependent pathways (22,72), effects on prostate-specific antigen levels, and association with prostate hyperplasia (73). Understanding the relationship between PFAS exposure and comorbidities, such as CVD, is particularly relevant because PFAS affects CVDs (74), and cardiovascular-related conditions are a major contributor to mortality among PCa patients (75). Although PFOS GM concentration in REPEL was 3.99 ng/mL, it is still higher than the threshold recommended by the American Heart Association for dyslipidemia screening using a lipid panel (>2 ng/mL) (48). This finding may have implications for clinical follow-up and preventive care.

### Lessons learned

This study demonstrated the feasibility of recruiting men with PCa, successfully enrolling 100 participants within 11 months of implementation. This was achieved through clear and frequent communication between the REPEL study team, oncology care providers, and biospecimen repositories and NJDOH laboratories (**Supplementary Figure 2**). The greatest challenges arose from volunteer tap water collections. Quality control issues were detected in seven kits due to inappropriate cold storage or shipping; however, the REPEL team worked with NJDOH to prepare a tutorial video and improve practices for collection, shipping, and handling to address these issues. Delivery of kits and collection of serum was often delayed due to missed appointments (28-33% of participants). Staffing changes on the REPEL team and at the laboratory during the study also required retraining and re-introduction of study procedures to colleagues at the hospital.

### Limitations and strengths

Our study had some limitations, including small sample size which limited our ability to conduct analyses of tap water and serum adjusted for covariates, and assess associations with health outcomes. Contemporary PFAS levels in water do not reflect historically high levels, and so the correlations between serum and tap water levels of PFAS may underestimate the true relationship between etiologically relevant tap water PFAS levels and serum levels. This study population was recruited in a department of urology and may overrepresent surgical and active surveillance patients, which may limit generalizability. Although the sample size was limited, this pilot supports the feasibility of accruing a larger cohort for future research. Additional study strengths include capture of PFAS sources through questionnaires, and collection of serum and tap water measures of PFAS to inform epidemiologic studies in cancer survivors. Integrating EHR will allow efficient passive capture of clinical factors including progression, treatment, and biomarkers.

## Conclusion

The REPEL cohort of men with PCa showed feasibility of collecting tap water samples and serum specimens to study health effects of PFAS in cancer survivors. This study is among the first to show a racial disparity in PFAS tap water and serum levels, with higher concentrations in non-White men with PCa. Serum concentrations of PFOS and PFOA were the highest among measured analytes, yet remained comparable to national and state estimates. Although PFAS were detected in participants’ tap water, correlations between serum and tap water remained modest. Future analyses will further clarify how PFAS impacts PCa and cardiometabolic health.

## Data Availability

The datasets generated during and/or analysed during the current study are available from the corresponding author on reasonable request.

## 8. Acknowledgments

We would like to express our sincere gratitude to our study participants for their support and time toward this research. Special thanks to the Urologic Oncology department at Rutgers Cancer Institute (Tina Mayer, Ryan Stephenson, Biren Saraiya, Ronald Ennis, Annie Daniel, Victorianna Belcastro, Vignesh Packiam, Danielle Velez Leitner), Biospecimen Repository and Histopathology Service at Rutgers Cancer Institute (Joseph Rosenberg), New Jersey Department of Health (David C. Riker, Sarah Desrochers, Shawn O’Leary, Diana Mathes, Anna Maria Marcel-Davila), Cancer Prevention and Outcomes Data Support at Rutgers Cancer Institute (Carolina Lozada), and the Cancer Prevention and Control Research Program at Rutgers Cancer Institute. We acknowledge and appreciate support from Barbara Weiland, Jimmie Staton, and Jerome C. Irons on study recruitment materials, questionnaires, and support with webinars for participants. We thank Jacob Hanna for support with data collection.

## 9. Author contributions

Conceptualization: HSI, SAJ, JEH, ESB, EVB; Data curation: SAJ, CO, JV, UI, SV, TLJ, SG, DG; Formal analysis: HSI, MP, MRS, TF, SO; Funding acquisition: HSI; Investigation: SAJ, HSI; Methodology: HSI, TF; Validation: SG, TLG, DG; Visualization: HSI, MRS; Writing – original draft: HSI, SAJ; Writing – review and editing: HSI, SAJ, CO, MRS, JV, JF, UI, SV, MP, TLJ, DG, SG, TF, SO, JMG, JEH, ESB, EVB.

## 11. Ethical Approval

This study was approved by the Rutgers University Institutional Review Board. Written informed consent was obtained from all participants prior to study participation.

