## Supplementary Materials for "Per- and Polyfluoroalkyl Substances Exposure in New Jersey Prostate Cancer Survivors: A Pilot Biomonitoring Study"

**Supplementary Methods**

**Supplementary Results**

**Supplementary Table 1: Comparison of sociodemographic and treatment characteristics of eligible vs non-eligible REPEL participants (N=207), 02/2025 to 03/2026**

**Supplementary Table 2: REPEL PFAS-containing Dietary and Beverage Frequency of Consumption, 115 participants, 02/2025 to 03/2026**

**Supplementary Table 3: REPEL residential tap water samples PFAS detects and concentrations (N=74)**

**Supplementary Table 4: Bivariate comparisons of tap water (N=74) and PFAS serum (N=63) levels of participants**

**Supplementary Table 5: REPEL serum samples PFAS detects and concentrations (N=63)**

**Supplementary Figure 1: REPEL data sources and study design**

**Supplementary Figure 2: REPEL stakeholders and collaborators**

**Supplementary Figure 3: Tap water collection kit materials**

**Supplementary Figure 4: REPEL study accrual at Rutgers Cancer Institute, New Brunswick, New Jersey, 02/2025 to 03/2026**

**Supplementary Figure 5: REPEL PFAS-containing Dietary and Beverage Consumption, 115 participants, 02/2025 to 03/2026**

**Supplementary Figure 6. Serum concentrations among REPEL participants and other population and highly PFAS exposed cohorts**

**Supplementary Methods**

*PFAS regulations in New Jersey*

New Jersey was chosen due to its history of drinking water PFAS contamination. Even when PFAS concentrations in drinking water are low, concentrations are magnified in humans with serum levels often as much as 100-times higher than in water (1,2). Furthermore, as certain PFAS have been phased out from consumer goods, water has become a more important contributor to exposure (1). New Jersey became the first state to develop regulatory levels for PFAS in drinking water with recommended MCLs for PFNA (13 ng/L) in 2015, PFOA (14 ng/L) in 2017, and PFOS (13 ng/L) in 2018 (3).

A study by Mueller et al. found the New Jersey Department of Environmental Protection (NJDEP) testing from 2019–2021 detected PFAS in 63% of community water systems, serving 84% of New Jersey’s population, showing that many New Jersey water systems continue to have detectable PFAS following regulatory actions (4). Compared to national estimates, New Jersey public water systems reported higher percentages of PFAS concentrations based on the analysis from the Third Unregulated Contaminant Monitoring Rule database (2013-2015) (5). Specifically, PFOA and PFNA concentrations ≥ 20 ng/L were observed in 10.2% vs. 1.9% and 2.3% vs. 0.2% of systems respectively, while PFOS levels ≥ 40 ng/L were identified in 3.4% vs. 1.9% nationwide (6). Notably, these findings are also reflected in biomonitoring data, as serum PFAS concentrations among New Jersey residents (2016-2018) exceeded those observed nationally in NHANES 2015-2016 (6). Moreover, because PFAS have long half-lives (2.7-8.5 years) (7), biomonitoring data may not yet reflect post-regulatory concentrations. Therefore New Jersey residents may still experience health impacts of exposure that occurred prior to the regulations (7,8).

*Stakeholder engagement*

The REPEL study engaged multiple stakeholders and collaborators at CINJ and New Jersey state government (**Supplementary Figure 2**). Beginning in July 2024 (8 months before study launch), the research team met with scientists at NJDOH to arrange protocols for shipping, processing and conducting PFAS assays for serum and tap water samples. In parallel, the REPEL research team met with staff at the Rutgers Cancer Health Equity Center of Excellence to solicit members of a CAB made of three citizen scientists. In addition to connections with state laboratory and CINJ community outreach programs, relationships were developed with CINJ shared resources for database development, and biospecimen repository and histopathology service. Members of the GU oncology team were contacted to discuss the study procedures and plan.

*Community advisory board*

The CAB was established in January 2025 in partnership with the Rutgers Cancer Health Equity Center of Excellence. Our research team met with Citizen Scientists twice (August 29, 2024 and September 12, 2024), who formed the CAB prior to study recruitment. The CAB provided feedback on the study brochure and fliers, as well as our consent form and questionnaire. After forming the CAB, the REPEL team met with the full group thrice (January 23, 2025, May 2, 2025, and September 22, 2025) to discuss progress and challenges with recruitment, and provide feedback on other dissemination materials (website posts, newsletters). These meetings helped to ensure that study materials minimized technical jargon and presented scientific information and study procedures clearly to eligible PCa patients.

Two webinars were held with study participants and moderated by CAB members (November 21, 2025 and March 19, 2026). During these webinars, the study team presented updates on recruitment, information about recent laboratory research on PFAS in New Jersey, and preliminary findings from the REPEL study regarding serum and tap water levels. Moderators prepared questions on behalf of the participants. All webinar recordings are available on our study website (9). To date, three newsletters have been published, two webinars were conducted, and study participants were emailed information of relevant studies. These activities have helped to enhance trust and transparency with study participants and their families and caregivers.

*Geographic data*

Residential addresses captured in the questionnaire were geocoded using a local geocoder built from street level business and residential data from New Jersey state records in ArcGIS Pro to protect privacy (10). Residential address histories were aggregated to counties before mapping to protect privacy. These address histories will be used to correlate historical geospatially-modeled PFAS levels and other environmental exposures prior to diagnosis to assess long-term impacts on progression and other chronic disease outcomes.

*Tap water samples and PFAS assessment*

Study participants received a tap water collection kit in-person at their upcoming scheduled appointment at CINJ. Materials can be viewed in **Supplementary Figure 3**. All materials were packed in a 12” x 10” x 9” inch insulated ice box by the REPEL study team. Each participant was given one 250 mL bottle pre-filled with reagent deionized water (highly purified and polished water- 18MΩ, <10 ppb Total Organic Compounds (TOCs)) and four empty bottles with ammonium acetate. Three bottles were designated for collecting tap water samples from home kitchen faucets, and the fourth was intended to be filled by the participant with the reagent serving as a control during shipment.

Participants were instructed to fill each of the three sample bottles to the shoulder. The kit also contained a pair of gloves, Ziploc® bags, and ice packs to prevent contamination and keep samples at an appropriate cold temperature of no less than 10°C during transport. Instructions, a tutorial video, data collections forms, and a prepaid return shipping label were included. The form asked participants to record the date and general time of collection, indicate whether samples were refrigerated after collection, and to confirm that all instructions were reviewed and followed.

Upon delivering the kit, our team informed the participants of the immediate steps needed to be taken after receiving the materials (such as placing the ice packs in the freezer and reagent water in the refrigerator); these instructions were also included in the box. After collection, samples were mailed to NJDOH-PHEL for PFAS analysis. Control samples were tested in parallel with experimental samples to identify any potential contamination during the transport process.

**Supplementary Results**

*Dietary exposures and water consumption*

PFAS exposure through food and drink sources were documented during the dietary habit section of the questionnaire. Frequencies can be viewed in **Supplementary Table 2** and **Supplementary Figure 5**. The most consumed (high frequency) beverages were water in plastic containers (69%), milk in plastic containers (48%), and other beverages in plastic containers (46%). The most consumed food was meat wrapped in plastic (70%). There was very low reported consumption of fast food, soda from fast food containers, or fish or shellfish.

Along with dietary habits, water consumption characteristics were also self-reported by participants (**Table 1**). Most men reported having a city/community/public water supply for their residential address (90%), while 10% had a private well. Most participants used filtered water as their main source of drinking water, either through filtered tap water (65%) or bottled water (23%), whereas unfiltered water (12%) was less commonly reported as their primary source. Among the 98 participants who reported drinking tap water, the median daily consumption was 32 oz (IQR: 8-48 oz) a day.

10.	Environmental Systems Research Institute. ArcGIS Pro. Redlands, CA: Esri; 2024.

**Supplementary Table 1: Comparison of sociodemographic and treatment characteristics of eligible vs non-eligible REPEL participants (N=207), 02/2025 to 03/2026**

|  | **Approached**^1^ | **REPEL**^2^ |
| --- | --- | --- |
| **N** | 83 | 124 |
| **Age at Diagnosis** | 67.00 (59.50, 72.00) | 64.00 (57.00, 68.00) |
| Unknown | 32 | 0 |
| **Race** |  |  |
| Asian | 10 (12%) | 8 (6%) |
| Black | 16 (19%) | 21 (17%) |
| White | 43 (52%) | 72 (58%) |
| Other | 14 (17%) | 23 (19%) |
| **Married** | 59 (71%) | 109 (88%) |
| Unknown | 2 (2%) | 1 (1%) |
| **Treatment Plan** |  |  |
| Active surveillance | 39 (50%) | 68 (57%) |
| Androgen deprivation therapy | 7 (9%) | 12 (10%) |
| Chemotherapy | 3 (4%) | 2 (2%) |
| Radiation | 4 (5%) | 2 (2%) |
| Surgery | 20 (26%) | 31 (26%) |
| No treatment plan | 5 (6%) | 4 (3%) |
| Unknown | 5 | 5 |

^1^ Includes participants who refused (n=51), were ineligible (n=12), requested additional time to make a decision (n=5), and withdrew (n=15).

^2^ Participants who consented to participate in the study.

**Supplementary Table 2: REPEL PFAS-containing Dietary and Beverage Frequency of Consumption, 115 participants, 02/2025 to 03/2026**

| **Diet Variable^1^** | **N (%)** |
| --- | --- |
| **N** | 115 |
| **Frequency of milk in plastic containers** |  |
| Never | 36 (31%) |
| Low | 24 (21%) |
| High | 55 (48%) |
| **Frequency of water in plastic containers** |  |
| Never | 5 (4%) |
| Low | 31 (27%) |
| High | 79 (69%) |
| **Frequency of drinks in plastic containers** |  |
| Never | 30 (26%) |
| Low | 32 (28%) |
| High | 53 (46%) |
| **Frequency of canned beverages** |  |
| Never | 28 (24%) |
| Low | 42 (37%) |
| High | 44 (38%) |
| Missing | 1 (1%) |
| **Frequency of canned foods** |  |
| Never | 16 (14%) |
| Low | 59 (51%) |
| High | 40 (35%) |
| **Frequency of meat wrapped in plastic** |  |
| Never | 13 (11%) |
| Low | 21 (18%) |
| High | 81 (70%) |
| **Frequency of cheese wrapped in plastic** |  |
| Never | 17 (15%) |
| Low | 36 (31%) |
| High | 61 (53%) |
| Missing | 1 (1%) |
| **Frequency of microwave popcorn** |  |
| Never | 81 (70%) |
| Low | 34 (30%) |
| **Frequency of ice cream** |  |
| Never | 24 (21%) |
| Low | 64 (56%) |
| High | 27 (23%) |
| **Frequency of tea using teabags** |  |
| Never | 47 (41%) |
| Low | 30 (26%) |
| High | 38 (33%) |
| **Frequency of fast food** |  |
| Never | 20 (17%) |
| Low | 77 (67%) |
| High | 18 (16%) |
| **Frequency of soda from fast food containers** |  |
| Never | 54 (47%) |
| Low | 48 (42%) |
| High | 13 (11%) |
| **Frequency of fish or shellfish** |  |
| Never | 12 (10%) |
| Low | 79 (69%) |
| High | 24 (21%) |

^1^Frequency of consumption was categorized as Never, Low, or High. Never indicates no reported consumption. Low includes responses of “a few times per year,” “once a month or less,” and “1 to 4 times a month.” High includes responses of “a few times a week,” “1 to 3 times a day,” and “more than 3 times a day.”

**Supplementary Table 3: REPEL residential tap water samples PFAS detects and concentrations (N=74)**

| **Analyte** | **N total** | **N detect** | **Percent detect** | **MDL (ng/L)** | **Water Concentration (ng/L)** | | | | | |
| --- | --- | --- | --- | --- | --- | --- | --- | --- | --- | --- |
|  |  |  |  |  | **Min** | **Q1** | **Median** | **Q3** | **Q90** | **Max** |
| 4:2FTS | 74 | 0 | 0 | 0.47 | <MDL | | | | | |
| 8:2FTS | 74 | 0 | 0 | 0.57 | <MDL | | | | | |
| 9Cl-PF3ONS | 74 | 0 | 0 | 0.48 | <MDL | | | | | |
| ADONA | 74 | 0 | 0 | 0.40 | <MDL | | | | | |
| NFDHA | 74 | 0 | 0 | 0.56 | <MDL | | | | | |
| PFDA | 74 | 0 | 0 | 0.47 | <MDL | | | | | |
| PFDoA | 74 | 0 | 0 | 0.59 | <MDL | | | | | |
| PFEESA | 74 | 0 | 0 | 0.28 | <MDL | | | | | |
| PFHpS | 74 | 0 | 0 | 0.82 | <MDL | | | | | |
| PFMBA | 74 | 0 | 0 | 0.38 | <MDL | | | | | |
| PFMPA | 74 | 0 | 0 | 0.40 | <MDL | | | | | |
| PFPeS | 74 | 0 | 0 | 0.41 | <MDL | | | | | |
| PFUnA | 74 | 0 | 0 | 0.49 | <MDL | | | | | |
| 11Cl-PF3OUdS | 74 | 2 | 2.7 | 0.55 | <MDL | | | | | |
| 6:2FTS | 73 | 3 | 4.1 | 0.37 | <MDL | | | | | |
| HFPO-DA | 74 | 11 | 14.9 | 0.44 | <MDL | | | | | |
| PFNA | 74 | 45 | 60.8 | 0.46 | 0.33 | 0.33 | 0.56 | 0.75 | 1.11 | 1.88 |
| PFHxS | 74 | 50 | 67.6 | 0.55 | 0.39 | 0.39 | 0.82 | 1.12 | 1.67 | 2.57 |
| PFOS | 74 | 54 | 73.0 | 0.66 | 0.46 | 0.46 | 2.29 | 2.89 | 4.52 | 7.17 |
| PFBS | 74 | 53 | 71.6 | 0.48 | 0.34 | 0.34 | 1.53 | 2.03 | 2.80 | 3.82 |
| PFHpA | 74 | 58 | 78.4 | 0.34 | 0.24 | 0.40 | 1.03 | 1.43 | 1.96 | 3.16 |
| PFPeA | 74 | 59 | 79.7 | 0.76 | 0.54 | 1.48 | 2.71 | 3.45 | 4.31 | 7.65 |
| PFHxA | 74 | 61 | 82.4 | 0.38 | 0.27 | 0.78 | 2.19 | 2.76 | 3.59 | 6.38 |
| PFOA | 74 | 63 | 85.1 | 0.38 | 0.27 | 1.21 | 3.75 | 5.27 | 8.04 | 12.50 |
| PFBA | 74 | 68 | 91.9 | 0.27 | 0.19 | 1.63 | 2.91 | 4.17 | 5.39 | 9.22 |

Abbreviation: MDL = Method detection limit

**Supplementary Table 4: Bivariate comparisons of tap water (N=74) and PFAS serum (N=63) levels of participants**

| ***Tap water (ng/L)*** | **PFHpS** | | | **PFHxS** | | | **PFNA** | | | **smPFOS** | | | **PFOA** | | | **PFOS** | | |
| --- | --- | --- | --- | --- | --- | --- | --- | --- | --- | --- | --- | --- | --- | --- | --- | --- | --- | --- |
|  | **GM** | **95% CI** | **P^1^** | **GM** | **95% CI** | **P^1^** | **GM** | **95% CI** | **P^1^** | **GM** | **95% CI** | **P^1^** | **GM** | **95% CI** | **P^1^** | **GM** | **95% CI** | **P^1^** |
| **Age (years)** |  |  |  |  |  |  |  |  |  |  |  |  |  |  |  |  |  |  |
| <65 |  |  |  | 0.81 | 0.70-0.94 | 0.474 | 0.56 | 0.48-0.64 | 0.667 |  |  |  | 2.93 | 2.08-4.12 | 0.187 | 1.71 | 1.33-2.21 | 0.686 |
| 65+ |  |  |  | 0.72 | 0.59-0.89 |  | 0.53 | 0.45-0.63 |  |  |  |  | 1.98 | 1.33-2.94 |  | 1.55 | 1.15-2.09 |  |
| **Race** |  |  |  |  |  |  |  |  |  |  |  |  |  |  |  |  |  |  |
| Non-white |  |  |  | 0.98 | 0.78-1.22 | 0.013 | 0.67 | 0.55-0.81 | 0.008 |  |  |  | 3.65 | 2.43-5.50 | 0.044 | 2.40 | 1.79-3.20 | 0.009 |
| White |  |  |  | 0.68 | 0.59-0.79 |  | 0.49 | 0.43-0.56 |  |  |  |  | 1.95 | 1.41-2.72 |  | 1.36 | 1.07-1.72 |  |
| **Risk group** |  |  |  |  |  |  |  |  |  |  |  |  |  |  |  |  |  |  |
| Low/intermediate |  |  |  | 0.74 | 0.63-0.87 | 0.555 | 0.55 | 0.48-0.63 | 0.747 |  |  |  | 2.22 | 1.60-3.08 | 0.529 | 1.58 | 1.24-2.01 | 0.714 |
| High/metastatic |  |  |  | 0.81 | 0.65-1.01 |  | 0.53 | 0.45-0.62 |  |  |  |  | 2.69 | 1.70-4.28 |  | 1.71 | 1.22-2.39 |  |
| ***Serum (ng/mL)*** | **PFHpS** | | | **PFHxS** | | | **PFNA** | | | **smPFOS** | | | **nPFOA** | | | **nPFOS** | | |
|  | **GM** | **95% CI** | **P^1^** | **GM** | **95% CI** | **P^1^** | **GM** | **95% CI** | **P^1^** | **GM** | **95% CI** | **P^1^** | **GM** | **95% CI** | **P^1^** | **GM** | **95% CI** | **P^1^** |
| **Age (years)** |  |  |  |  |  |  |  |  |  |  |  |  |  |  |  |  |  |  |
| <65 | 0.10 | 0.07–0.14 | 0.816 | 1.19 | 0.81–1.73 | 0.889 | 0.36 | 0.27–0.48 | 0.811 | 1.91 | 1.22–2.99 | 0.643 | 1.32 | 1.01–1.73 | 0.924 | 2.30 | 1.58–3.37 | 0.584 |
| 65+ | 0.09 | 0.06–0.14 |  | 1.14 | 0.79–1.66 |  | 0.34 | 0.25–0.46 |  | 1.65 | 1.08–2.52 |  | 1.35 | 1.02–1.78 |  | 1.98 | 1.34–2.92 |  |
| **Race** |  |  |  |  |  |  |  |  |  |  |  |  |  |  |  |  |  |  |
| Non-white | 0.16 | 0.12–0.20 | 0.019 | 1.61 | 1.37–1.89 | 0.102 | 0.53 | 0.41–0.68 | 0.007 | 3.00 | 2.38–3.78 | 0.019 | 1.70 | 1.40–2.06 | 0.095 | 3.74 | 3.04–4.60 | 0.004 |
| White | 0.08 | 0.05–0.11 |  | 1.00 | 0.69–1.45 |  | 0.29 | 0.22–0.38 |  | 1.38 | 0.91–2.09 |  | 1.20 | 0.92–1.55 |  | 1.63 | 1.14–2.34 |  |
| **Risk group** |  |  |  |  |  |  |  |  |  |  |  |  |  |  |  |  |  |  |
| Low/intermediate | 0.09 | 0.06-0.13 | 0.300 | 1.02 | 0.70-1.48 | 0.170 | 0.31 | 0.24-0.41 | 0.132 | 1.54 | 0.99-2.38 | 0.220 | 1.21 | 0.93-1.57 | 0.159 | 1.83 | 1.27-2.64 | 0.133 |
| High/metastatic | 0.12 | 0.08-0.17 |  | 1.51 | 1.18-1.94 |  | 0.44 | 0.33-0.58 |  | 2.32 | 1.76-3.05 |  | 1.63 | 1.30-2.04 |  | 2.85 | 2.04-3.98 |  |

Abbreviation: GM=geometric mean. ^1^P-for-heterogeneity in geometric means comparing serum or tap water PFAS across levels of age, race and risk group.

**Supplementary Table 5: REPEL serum samples PFAS detects and concentrations (N=63)**

| **Analyte** | **N total** | **N detect** | **Percent detect** | **LOD (ng/mL)** | **Serum Concentration (ng/mL)** | | | | | |
| --- | --- | --- | --- | --- | --- | --- | --- | --- | --- | --- |
|  |  |  |  |  | **Min** | **Q1** | **Median** | **Q3** | **Q90** | **Max** |
| ADONA | 63 | 0 | 0 | 0.01 | <LOD | | | | | |
| HFPO-DA | 63 | 0 | 0 | 0.01 | <LOD | | | | | |
| Et-PFOSA-AcOH | 63 | 3 | 4.8 | 0.01 | <LOD | | | | | |
| 9Cl-PF3ONS | 63 | 4 | 6.3 | 0.01 | <LOD | | | | | |
| PFDoA | 63 | 7 | 11.1 | 0.02 | <LOD | | | | | |
| PFHxA | 63 | 7 | 11.1 | 0.02 | <LOD | | | | | |
| PFBuS | 63 | 8 | 12.7 | 0.01 | <LOD | | | | | |
| Sb-PFOA | 63 | 11 | 17.5 | 0.03 | <LOD | | | | | |
| Me-PFOSA-AcOH | 63 | 24 | 38.1 | 0.01 | <LOD | | | | | |
| PFHpA | 63 | 25 | 39.7 | 0.03 | <LOD | | | | | |
| PFOSA | 63 | 32 | 50.8 | 0.01 | 0.00 | 0.00 | 0.00 | 0.01 | 0.02 | 0.09 |
| PFUnA | 63 | 41 | 65.1 | 0.01 | 0.01 | 0.01 | 0.05 | 0.12 | 0.19 | 1.43 |
| PFDeA | 63 | 45 | 71.4 | 0.03 | 0.02 | 0.02 | 0.07 | 0.11 | 0.23 | 1.98 |
| PFHpS | 63 | 60 | 95.2 | 0.01 | 0.00 | 0.07 | 0.13 | 0.20 | 0.26 | 0.42 |
| PFHxS | 63 | 62 | 98.4 | 0.01 | 0.01 | 0.97 | 1.40 | 1.98 | 2.99 | 6.04 |
| PFNA | 63 | 62 | 98.4 | 0.02 | 0.01 | 0.23 | 0.39 | 0.56 | 0.74 | 1.79 |
| smPFOS | 63 | 62 | 98.4 | 0.01 | 0.01 | 1.62 | 2.10 | 3.28 | 4.62 | 12.30 |
| nPFOS | 63 | 61 | 96.8 | 0.05 | 0.04 | 1.54 | 2.55 | 3.82 | 5.22 | 30.70 |
| nPFOA | 63 | 63 | 100 | 0.02 | 0.06 | 1.06 | 1.55 | 1.98 | 2.62 | 5.90 |
| PFOA^1^ | 63 | - | - | - | 0.07 | 1.08 | 1.57 | 2.01 | 2.64 | 5.92 |
| PFOS^2^ | 63 | - | - | - | 0.04 | 3.40 | 4.82 | 7.32 | 10.12 | 35.19 |

Legend: LOD= Limit of detection, ^1^Sum of Sb-PFOA and n-PFOA, ^2^Sum of smPFOS and nPFOS

**Supplementary Figure 1: REPEL data sources and study design**

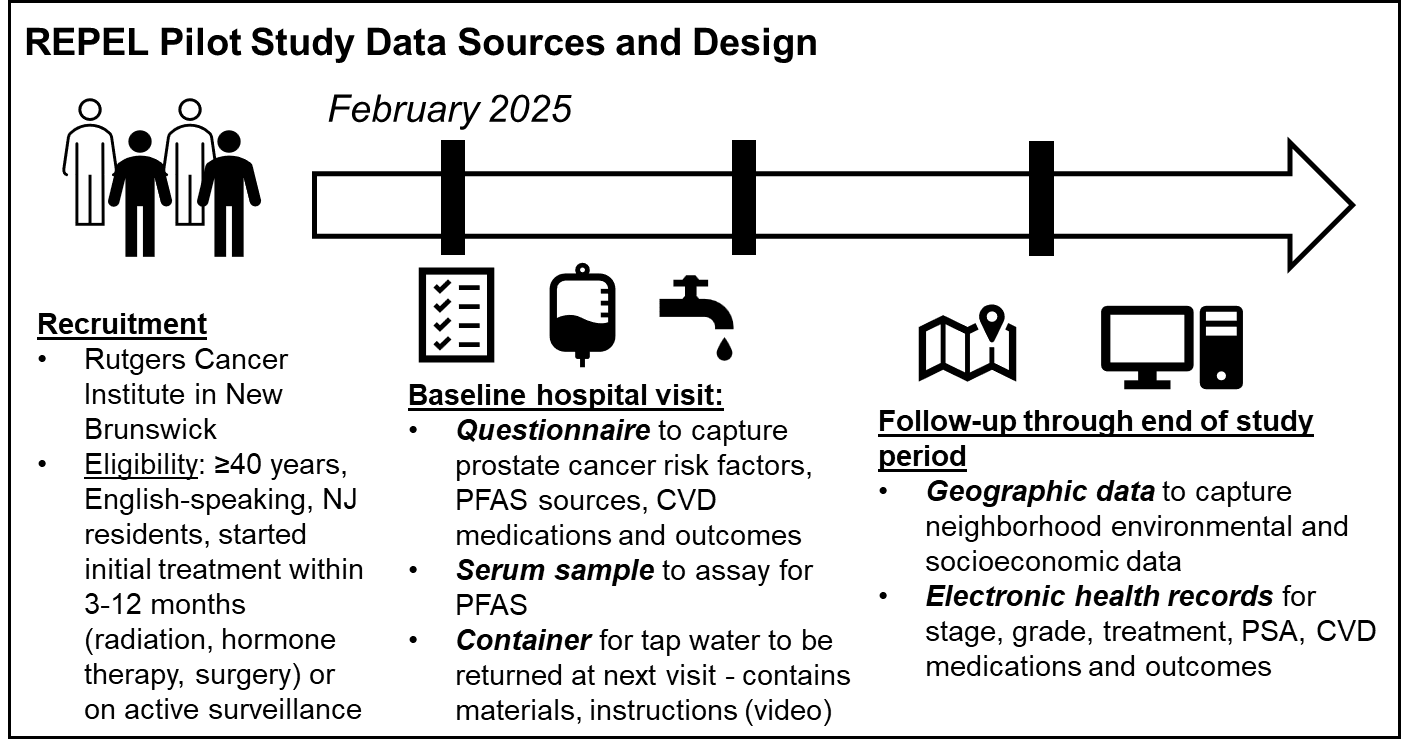

**Supplementary Figure 2: REPEL stakeholders and collaborators**

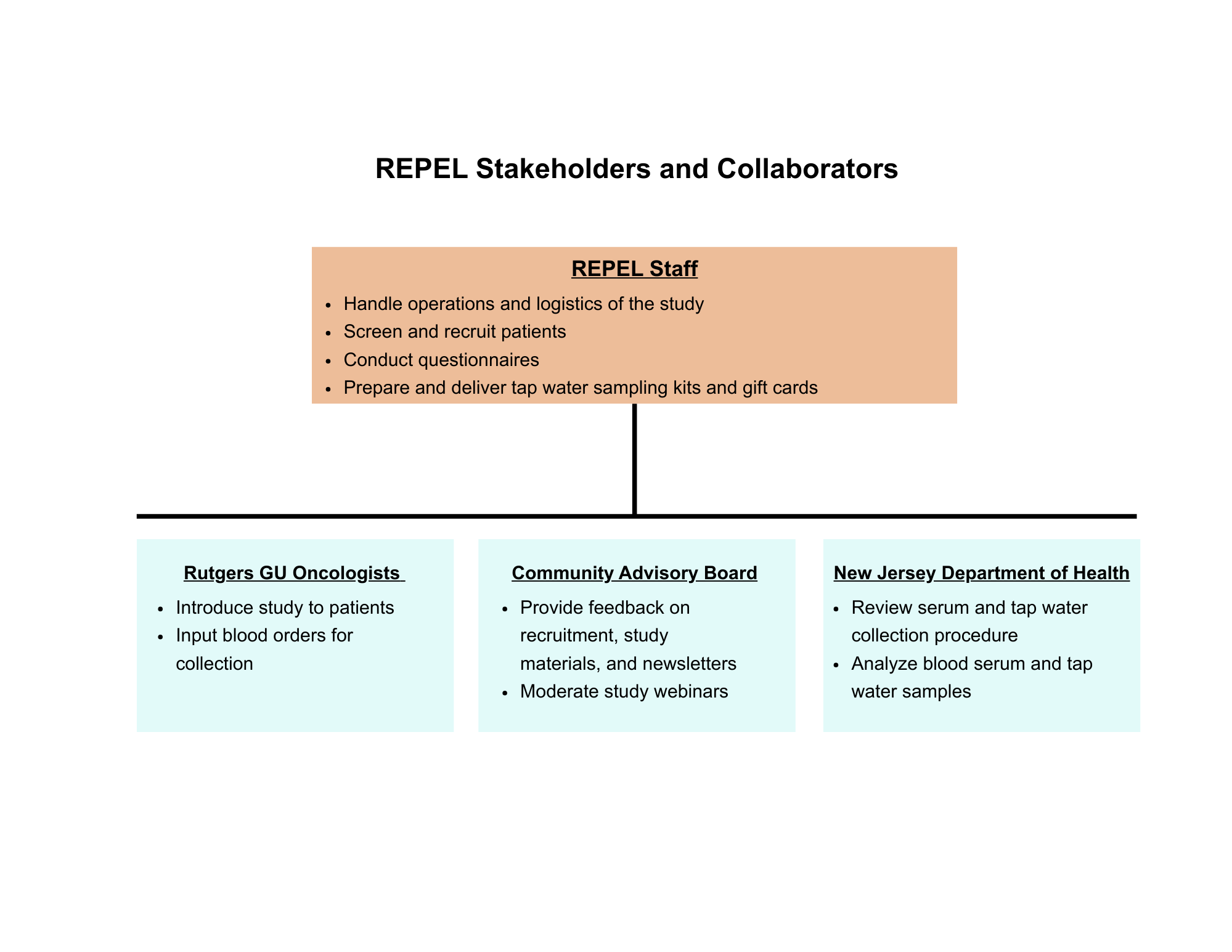

**Supplementary Figure 3: Tap water collection kit materials**

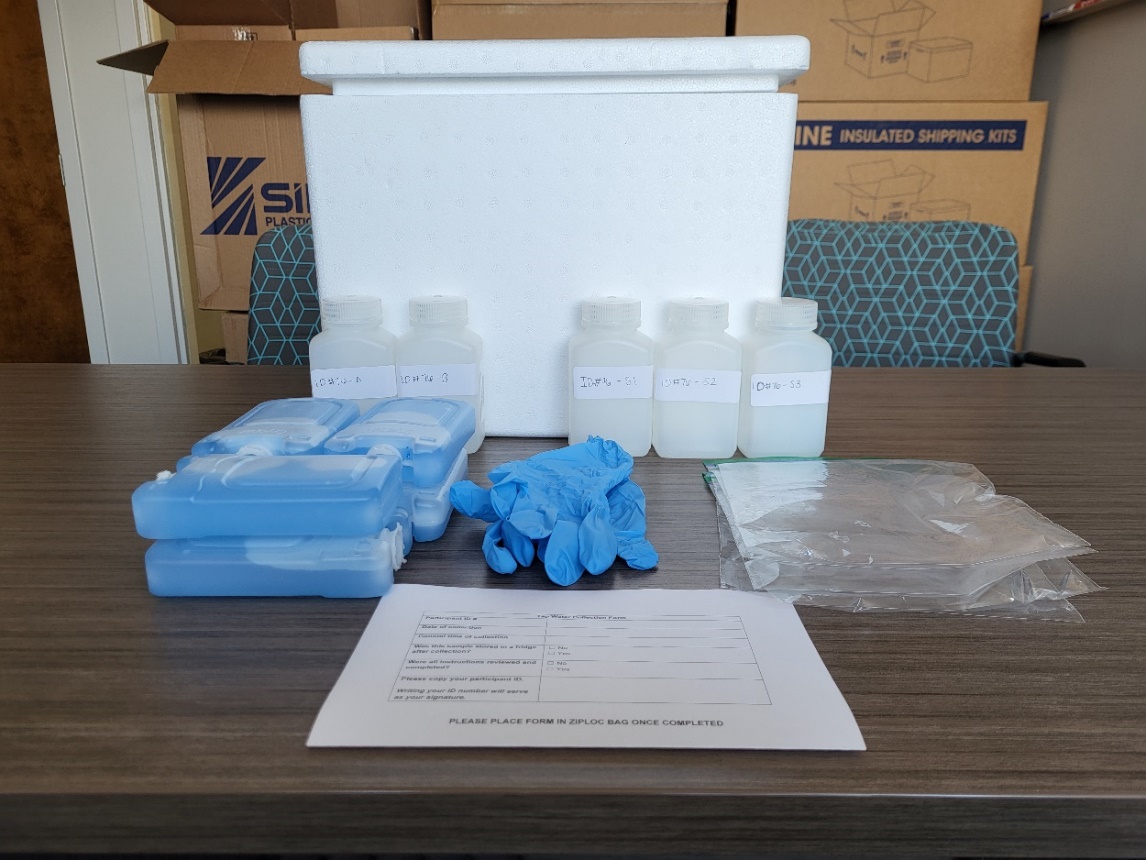

**g**

**f**

**c**

**b**

**d**

**e**

**a**

Legend: a. 6 Ice packs, b.1 pair of gloves, c. 5 Ziploc® bags, d. 3 plastic bottles labeled “S1”, “S2”, and “S3”, e. 2 plastic bottles labeled “A” (pre-filled with water) and “B” (contains symbol on the bottle lid), f. Tap water collection form and shipping label, g. Ice box

**Supplementary Figure 4: REPEL study accrual at Rutgers Cancer Institute, New Brunswick, New Jersey, 02/2025 to 03/2026**

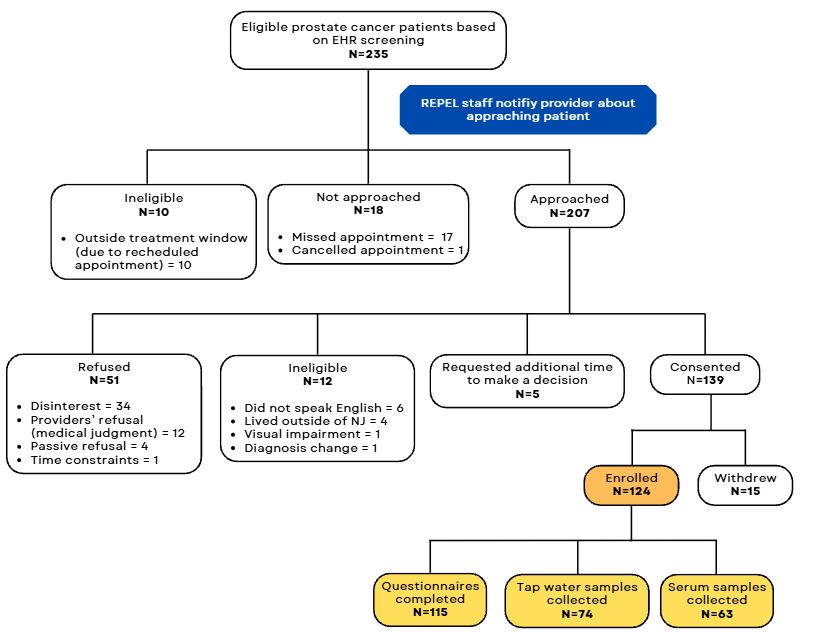

Legend: Abbreviations: EHR=Electronic health record, NJ = New Jersey

**Supplementary Figure 5: REPEL PFAS-containing Dietary and Beverage Consumption, 115 participants, 02/2025 to 03/2026**

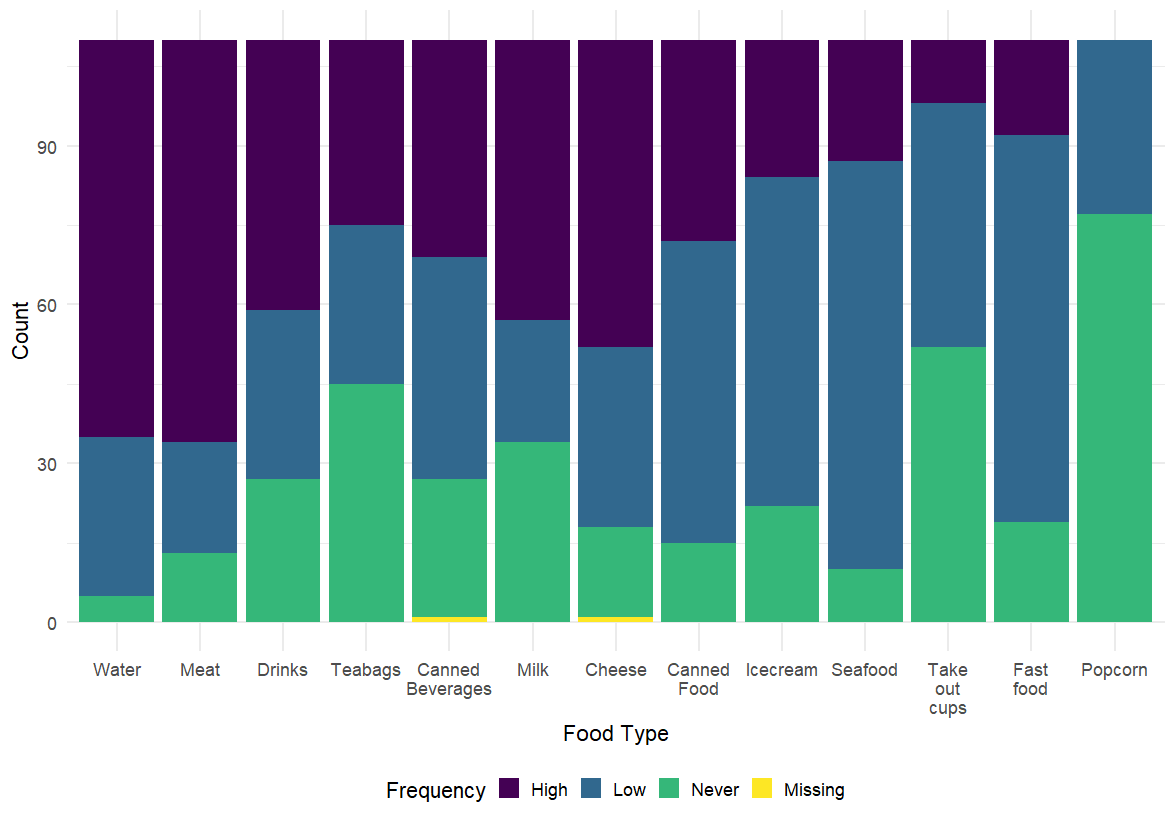

Legend: Frequency of consumption was categorized as Never, Low, or High. Never indicates no reported consumption. Low includes responses of “a few times per year,” “once a month or less,” and “1 to 4 times a month.” High includes responses of “a few times a week,” “1 to 3 times a day,” and “more than 3 times a day.

**Supplementary Figure 6. Serum concentrations among REPEL participants and other population and highly PFAS exposed cohorts**

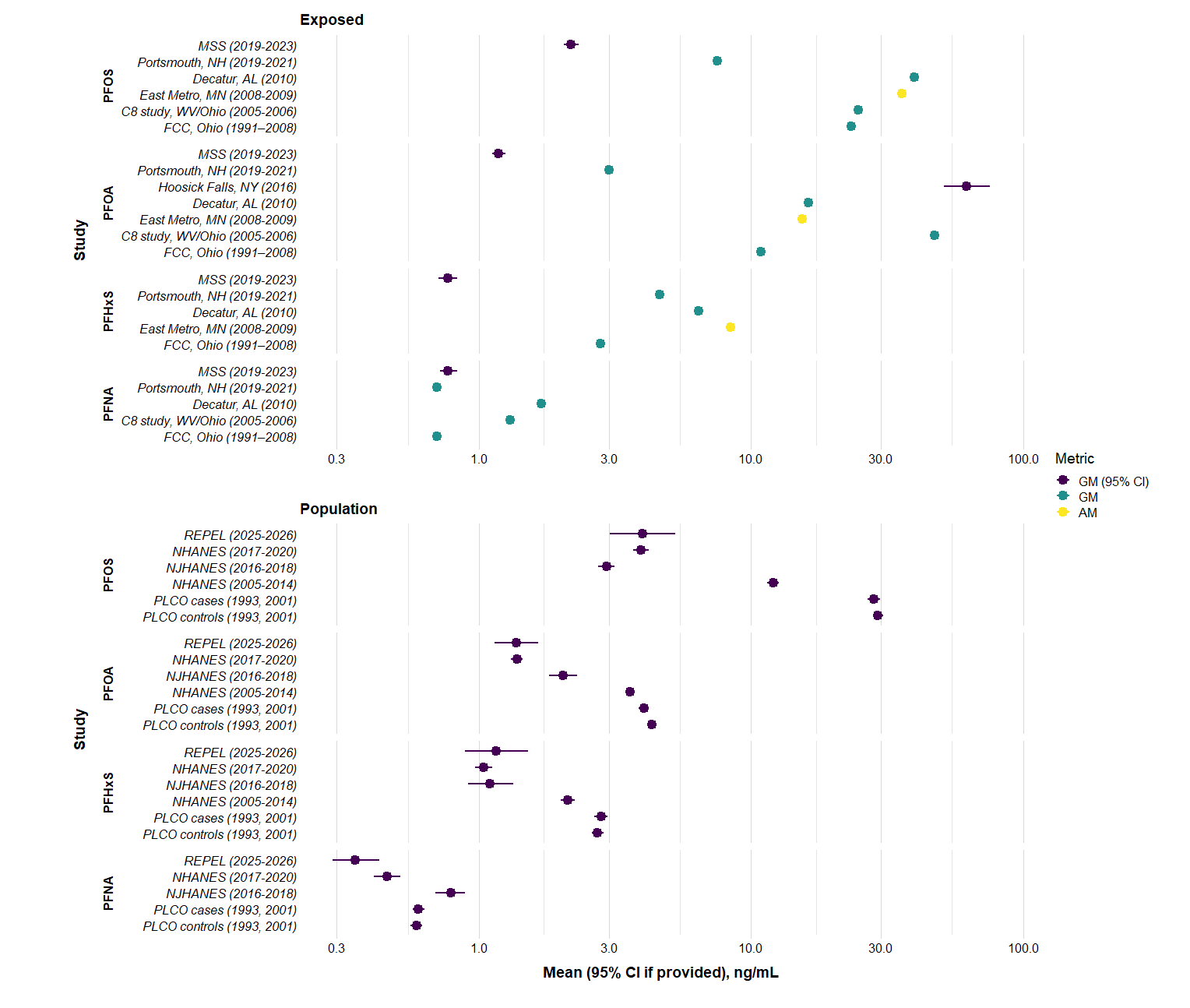
